# A scoping review of definitions and conceptualizations of ecosyndemics in syndemic research

**DOI:** 10.1101/2025.07.24.25331762

**Authors:** Yassir Gnaoui, M. Nienke Slagboom, Jessica C. Kiefte-de Jong, Mariëlle A. Beenackers

## Abstract

**Background:** Ecosyndemic research focuses on the role of environmental contextual factors in syndemics. However, there is no consensus on the difference between syndemics and ecosyndemics, as the physical environment is often also included in syndemic studies. This review aims to describe the definitions used, environmental aspects included, and methods employed in ecosyndemic and syndemic studies with an environmental contextual focus.

**Methods:** We conducted a literature search across five databases. We screened articles in two phases: software-assisted screening and manual full-text screening. To distinguish common elements, ecosyndemic and syndemics definitions were coded. Information regarding methods, factors related to the physical environment, and study results were extracted from the empirical articles.

**Results:** A total of 169 publications were included, 16 used an ecosyndemic definition, 137 a syndemic one, and 16 a global syndemic definition. Most articles were nonempirical (n=112). Two types of ecosyndemic conceptualizations were distinguished: the extension conceptualization (n=10), which emphasizes the role of the physical environmental context in combination with other contextual factors, and the trigger conceptualization (n=6), which focuses on how environmental factors can trigger a syndemic. The built environment was studied most frequently (n=48), the food environment the least (n=18).

**Conclusion:** The ecosyndemic framework highlights the role of the physical and social environment in worsened population-level health outcomes, which could inform health policies. Ecosyndemic research should focus on a multidisciplinary understanding of it through knowledge triangulation derived from different scientific disciplines to ensure that all aspects of ecosyndemics—clustering, interaction, and social and environmental contextual drivers—are included.

## 1. Introduction

Environmental contextual drivers of health are increasingly gaining academic attention. In this review we focused on ecosyndemics, which are a specific type of syndemic that focuses on the influence of the environment. Syndemic research focuses on the clustering and interaction of multiple diseases, with particular attention to the contextual factors that contribute to disease clustering within a population and a given context (Singer, 1996). Since the first use of syndemic theory—to conceptualize the clustering and adverse interaction of substance abuse, violence, and HIV/AIDS—the physical environment has been recognized as a potential contextual driver of disease clustering and interaction (Singer, 1996).

Syndemics research is structured around three core criteria: 1) two (or more) diseases cluster in a specific population in time or place, 2) the clustering of diseases results in synergistic interactions that affect the affected populations’ health burdens, and 3) contextual factors drive this disease clustering and interaction. This third criterion, context, is especially relevant to the growing body of ecosyndemic research, which examines environmental conditions as drivers of poor health outcomes. However, there has been no consensus on how to integrate and operationalize the physical environment within syndemic studies.

Although the role of the physical environment within syndemic theory is not conclusively defined, at the population level, all-cause mortality rates are associated with adverse living environments (Vandeninden et al., 2024). The environmental context influences population-level health outcomes through disease–environment interactions (Singer, 2016) and interactions with other contextual drivers of syndemics (Singer 2010a). An example of this is how the construction of the Belo Monte dam in Brazil led to ecological changes such as altered waterways that led to an increase in mosquitoes and vector-borne diseases (Tallman et al., 2022). The influx of male laborers was also linked to an increase in sex-work and sexually-transmitted infections. The contextual synergies observed during and after the construction of the Belo Monte dam led to disease–disease interactions such as the interaction in which schistosomiasis—a vector borne disease—can lead to greater susceptibility to human papillomavirus infection and subsequently a greater susceptibility to sexually transmitted diseases such as HIV (Tallman et al., 2022). Syndemics need to be understood from the perspective of the local context (Weaver & Kaiser, 2022). This includes the various localized physical environmental aspects that interact with the social context and impact disease clustering and adverse disease interaction (Rhodes et al., 2005). Therefore, to advance our understanding of how the physical environment affects syndemic health outcomes, it is essential to further distinguish how the physical environment is integrated within ecosyndemic and syndemic studies with an environmental contextual focus.

As an addition to syndemic theory, the term “ecosyndemic” was introduced to investigate disease and biosocial interactions that are driven by environmental factors such as climate change (Baer & Singer, 2009) and altered physical environments (Bare & Singer, 2009; Singer, 2025). The initial use of ecosyndemics focused on environmental changes caused by human actions (Baer & Singer, 2009). Since Baer and Singer introduced ecosyndemics in 2009, a growing number of studies have used the definition and have focused on how the physical environmental context drives disease clustering and interaction. Consequently, with the expansion of the field, new methodological questions have arisen regarding the definition and conceptualization of ecosyndemics as well as the implementation of environmental factors in ecosyndemic and syndemics studies. In this review, we examine the definitions and methods used within ecosyndemic and syndemic studies with an environmental contextual focus (from here on referred to as syndemic studies).

Although the ecosyndemic concept implies that environmental factors have a considerable influence on disease clustering and adverse disease interactions, the extent to which the physical environment plays a role in traditional syndemics is less clear. This makes it more difficult to distinguish between ecosyndemic and syndemic studies.

### 1.1. The conceptualization of ecosyndemics

We hypothesized that the ecosyndemic framework can be divided into two conceptualizations (Figure 1). In the first, ecosyndemics are an extension of syndemics. Within this conceptualization, ecosyndemics are similar to syndemics but have an added environmental component attached. This conceptualization can be interpreted as an added fourth criterion on top of the three syndemic criteria of clustering, interaction, and context. This is in contrast to traditional syndemic research where the role of socioeconomic contextual factors are emphasized (Gizamba et al., 2023; Pirrone et al., 2021). The extension conceptualization of ecosyndemics emphasizes the effect of environmental factors and/or environmental change on syndemics as an added layer of complexity. In addition, when viewed from a temporal perspective, environmental factors have a more long-term effect on the syndemic in a way that is comparable to how contextual factors are described in the third syndemic criterion as drivers of disease clustering and synergetic disease interactions. The second conceptualization defines ecosyndemics as syndemics that are triggered by environmental change or factors. Within this “trigger” conceptualization, the emphasis lies on sudden changes and alterations in physical environments that set a syndemic in motion. Compared to the extension conceptualization, the environmental changes are often more abrupt and disruptive.

**Figure 1.**
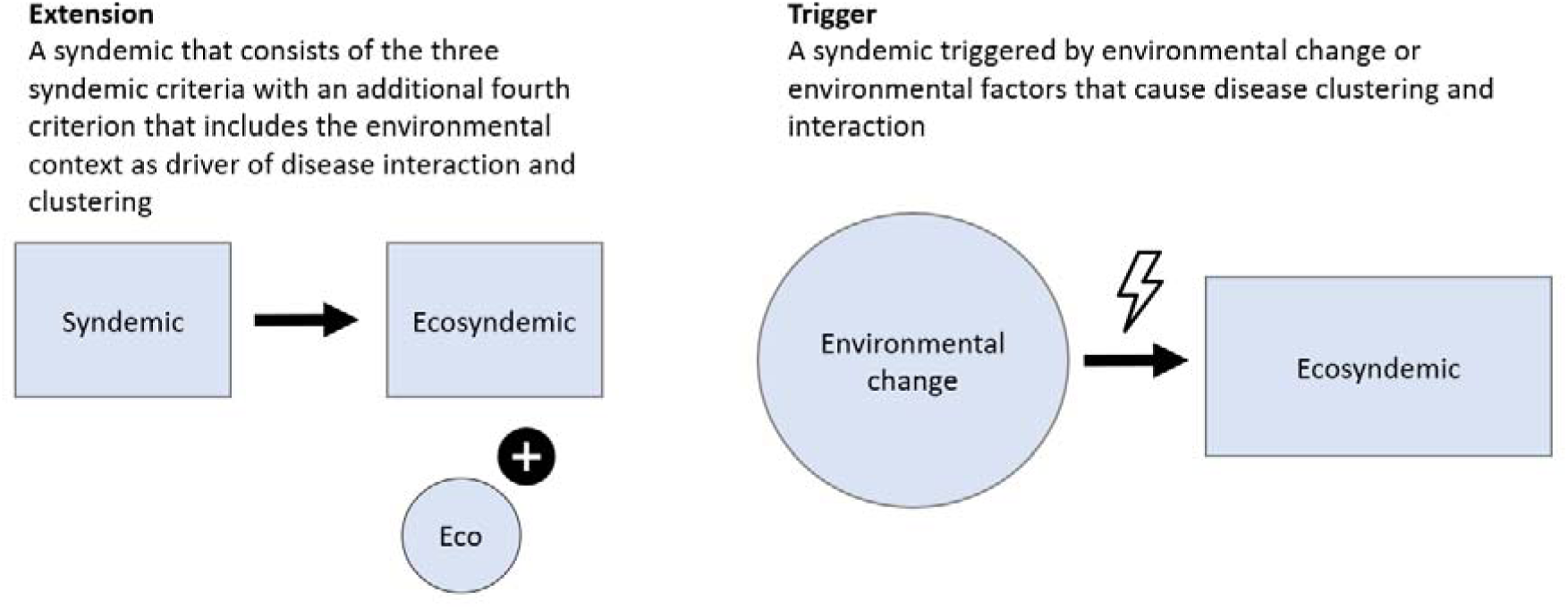
Visual depiction of the extension and trigger conceptualizations of ecosyndemics.

### 1.2. The role of the physical environment in syndemic theory

Earlier reviews have investigated the role of context and place in syndemic theory. Pirrone et al. (2021) found that the contextual scope of syndemics is often focused on the micro context, such as living conditions (e.g., housing and neighborhood), and socioeconomic indicators, such as income and education. In addition, challenges exist in connecting micro-level with macro-level contextual factors, such as political and societal characteristics and environmental processes (Pirrone et al., 2021). More recently, Gizamba et al. (2023) investigated the integration of contextual factors in syndemic studies and distinguished two different aspects of place:: ‘the social condition of a place’ and ‘the physical condition of a place’ in which the social condition of a place refers to factors such as income inequality, violence, and experience of stigma. The physical condition of a place refers to factors such as poor housing conditions, rurality or urbanicity, and air quality (Gizamba et al., 2023). The majority of studies have focused on the social condition of place, and contextual aspects have not been further categorized into micro-, meso-, and macro-level factors (Gizamba et al., 2023). No review has yet focused on the integration of the physical environment within syndemic studies or the methods used in ecosyndemic and syndemic studies.

### 1.3. Aims

This review aims to create an overview of the integration of environmental factors and the definitions used in ecosyndemic and syndemic studies. It therefore describes 1) the different ecosyndemic definitions and the definitions used in syndemic studies, 2) the environmental aspects included in syndemic studies as well as how these aspects are implemented in research, and 3) the methods used to study ecosyndemics and syndemics.

## 2. Methodology

To investigate the integration of environmental factors and the definitions used in ecosyndemic and syndemic studies, a scoping review was conducted. A scoping design was chosen in line with the aims of this review: We did not want to create an exhaustive listing of all the studies. Instead, the aim was to guide future ecosyndemic research by creating an overview of the predominantly used ecosyndemic definitions and the methods for integrating environmental factors.

### 2.1. Search strategy

A two-part search query was employed to include articles that used the term “ecosyndemics” and “syndemics” with an environmental contextual focus. This resulted in a search term that included articles that contained the term “ecosyndemics” or “syndemics” with the addition of a term related to the physical environment (Appendix A). After the first-phase screening, a hand search was conducted on the reference lists of the included articles that explicitly mentioned ecosyndemics. These articles were retrieved using the first component of the search term. Because there was a key ecosyndemic publication (Singer, 2025) during the finalization of this review, it was included, and a second hand search was conducted using this publication’s reference list.

### 2.2. Inclusion and exclusion criteria

Peer-reviewed articles and book chapters that reported on ecosyndemics or syndemics with an environmental contextual focus were considered for screening. Other types of publications, such as conference proceedings, dissertations, and theses, were excluded. Articles that focused on animal health were also excluded.

Using ASReview, publication titles and abstracts were screened according to the following eligibility criteria: The study 1) has an ecosyndemic or syndemic scope, 2) focuses on human health conditions, and 3) focuses on the effects of the physical environment on health. In the second phase, the included papers were exported from ASReview and then the full text screened based on the following eligibility criteria: The study includes 1) a definition of ecosyndemics or syndemics, 2) aspects related to disease clustering, and 3) the impact of the physical environment on the syndemic. ASReview is an open-source machine learning-aided pipeline with an active learning tool that can speed up the screening process (van de Schoot et al., 2021).

### 2.3. Study screening and extraction

The search results were downloaded in.ris format and imported into Endnote 21. Duplicates were removed using the duplicate tool in Endnote. The remaining papers were then imported into ASReview LAB Oracle Version 1.2.1 for first-phase screening.

ASReview first creates a training set through labeled records that are provided by the reviewer. Following from this, the active learning cycle starts. During this cycle, a classification algorithm is trained on the labeled records using a balancing strategy to deal with the data being unbalanced, and relevance scores are estimated for all unlabeled records. Then the electronic learning assistant chooses a record to show, and the reviewer then screens it, labeling it as relevant or irrelevant, after which the newly labeled record is moved to the training data set and the cycle starts again (ASReview, 2022). To make relevance predictions, we selected Naïve Bayes as the classifier in ASReview, and for the feature extraction technique, we selected the term frequency-inverse document frequency (TF-IDF). TF-IDF is a feature extraction technique that estimates how relevant a word is to a certain category of documents by considering the word or term frequency (TF) in the document and how unique or infrequent (IDF) a word is in the full body of work; higher values are assigned to topic representative words, while common words are devalued through TF-IDF (Zhang et al., 2020). Words are regarded as representative and are kept if they have higher TF-IDF weights, while those with lower weights are regarded as less representative and are discarded (Zhang et al., 2020). The use of the Naïve Bayesian classification in combination with a TF-IDF feature extraction technique can reduce the required number of screened publications by 71.7% at the cost of losing 5% of relevant publications and is one of the best-performing models for screening prioritization (Ferdinands et al., 2023). Human reviewers usually fail to identify 10% of the relevant papers (Wang et al., 2020), ASReview aims to find 95% of the relevant publications (ASReview, 2022). In a study by Schoot et al. (2021) results showed that 95% of eligible studies will be found in ASReview after screening 8% to 33%. In line with these results a threshold of 35% was maintained to aim for an inclusion of the majority of relevant publications.

## 3. Data processing

Two data extraction sheets were created in Microsoft Excel. Separate sheets were created for nonempirical and empirical articles. The nonempirical extraction sheet included elements regarding the article title, author, and aim; the definition of ecosyndemic; and the citation(s) used in the ecosyndemic definition. The definitions of ecosyndemics were thematically coded to distinguish between which definitions are used within the field and how.

The extraction sheet that was used for the empirical articles consisted of the following elements: article title, author, and aim; citation of ecosyndemic or syndemic definition; type of study; environmental variable; dependent variable; sample; methods; results; and conclusion. The environmental variables were categorized using the exposome framework (Beulens et al., 2022), which divides environmental drivers of disease into four separate categories: the built environment, described as the physical space in which we live and work; the social environment, described as the social relationships and social context in which groups of people live and interact; the physico-chemical environment, consisting of the chemical or physical agents present in our local area; and the lifestyle/food environment, which comprises the accessibility, availability, affordability, and promotion of foods and food retailers in a neighborhood. Because this review focused on the physical environment, the social environment was dropped as a category, and the ecological environment was added as a fourth category, which included factors related to ecological processes, such as climate change, and natural hazards, such as flooding. The ecological environment is included as a separate factor because a substantial part of ecosyndemic studies have focused on the effects of climate change and natural disasters on disease clustering and interaction.

Following data extraction, the syndemic and ecosyndemic definitions were coded according to the syndemic criteria. The primary categories used were the three main syndemic criteria: type of clustering, type of interaction, and type of contextual factors. The contextual factors were categorized according to the Context and Implementation of Complex Interventions (CICI) framework (Pfadenhauer et al., 2017), which consists of seven factors: geographical, epidemiological, sociocultural, socioeconomic, political, legal, and ethical.

Because clustering and interaction type were less suited to categorization according to frameworks, these criteria were coded inductively. In addition to the syndemic criteria, the following individual categories were included: definition, population (level), and outcome. Coding was conducted in Atlas.ti 24.

For the empirical studies, the quantitative studies were classified as longitudinal if the analysis included a time component or if the authors described the study as longitudinal. Studies were identified as cross-sectional pooled if the data were collected from multiple time points but analyzed cross-sectionally, meaning that no time component was included in the analysis. Studies that analyzed data from only one time point were considered cross-sectional. For mixed studies, the quantitative component was identified using the same categories that were used for the quantitative studies.

## 4. Results

### 4.1. Study selection

A literature search was conducted on work that was published before November 15, 2023. During the initial search, 5,184 publications were retrieved, of which 2,897 were duplicates. Screening using ASReview was conducted on 2,287 publications, of which 563 were manually excluded, and 1,488 were excluded through ASReview’s automated selection process, resulting in a total of 236 selected publications. The hand search yielded 24 publications, which were then added to the 236 obtained from the previous screening. Three articles obtained from the hand search were duplicates, and three were not retrieved. Of the 254 publications that were obtained for full-text screening, 90 were excluded. A chapter on ecosyndemics was published in a book on the anthropology of human and planetary health (Singer, 2025) during the finalization of this review. This publication was included and a second hand search conducted using its reference list, which yielded five additional publications: four nonempirical and one empirical study. This resulted in the final inclusion of 169 publications, of which 112 were nonempirical studies and 57 were empirical.

**Figure 2.**
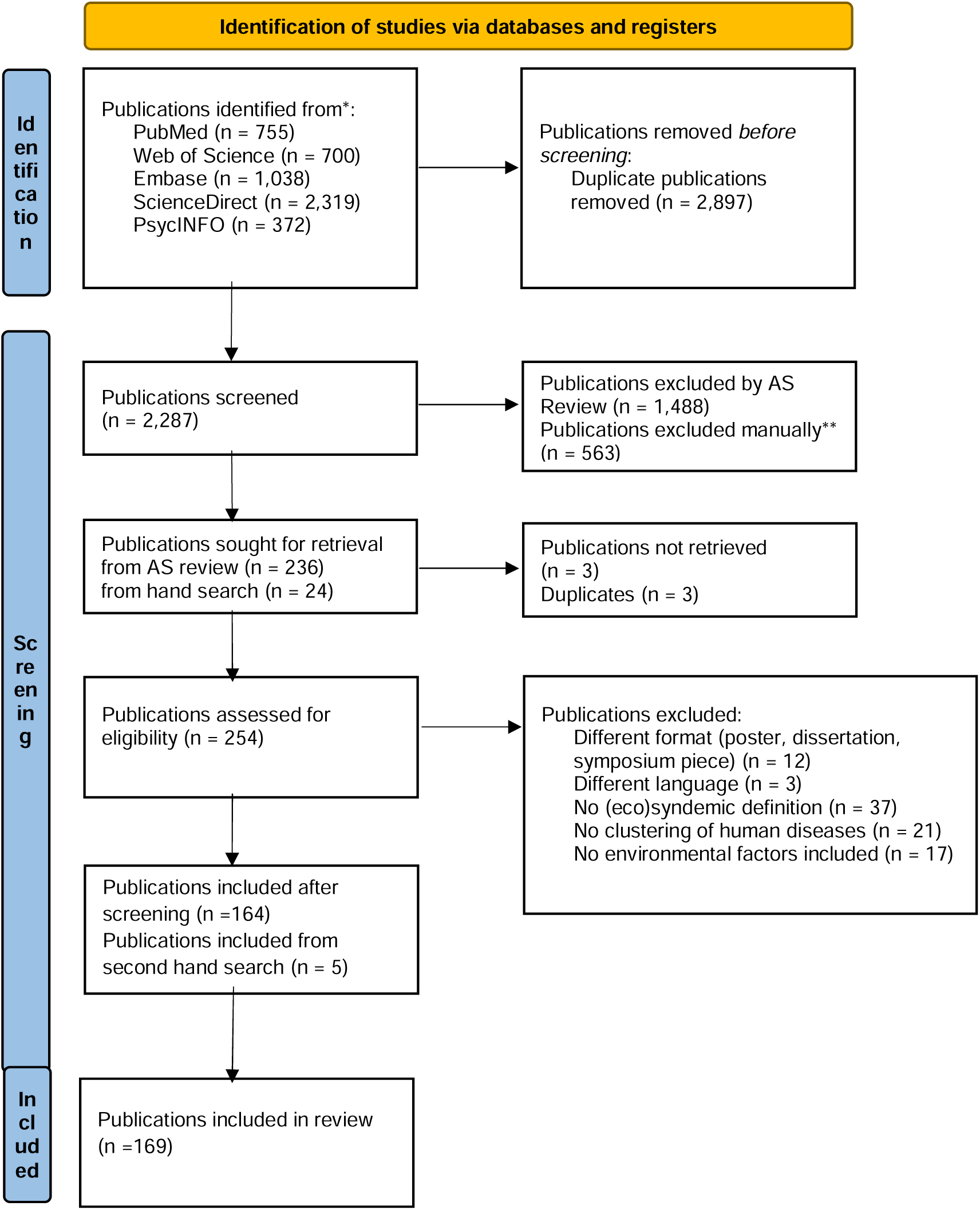
Prisma Flowchart of Screening and Selection Process.

### 4.2 Ecosyndemic definitions

Table 1 shows the unique definitions of ecosyndemics found in the publications (n = 16). The ecosyndemic definitions were grouped according to the “extension” and “trigger” conceptualizations: 1) ecosyndemics as an extension of syndemics, in which environmental factors play an essential role as drivers of disease clustering and adverse disease interactions in addition to adverse social contextual factors, and 2) the trigger conceptualization of ecosyndemics, in which environmental factors/changes act as a trigger that sets a syndemic in motion. Ten studies used the extension conceptualizations, and six drew on the trigger conceptualization.

**Table 1.**
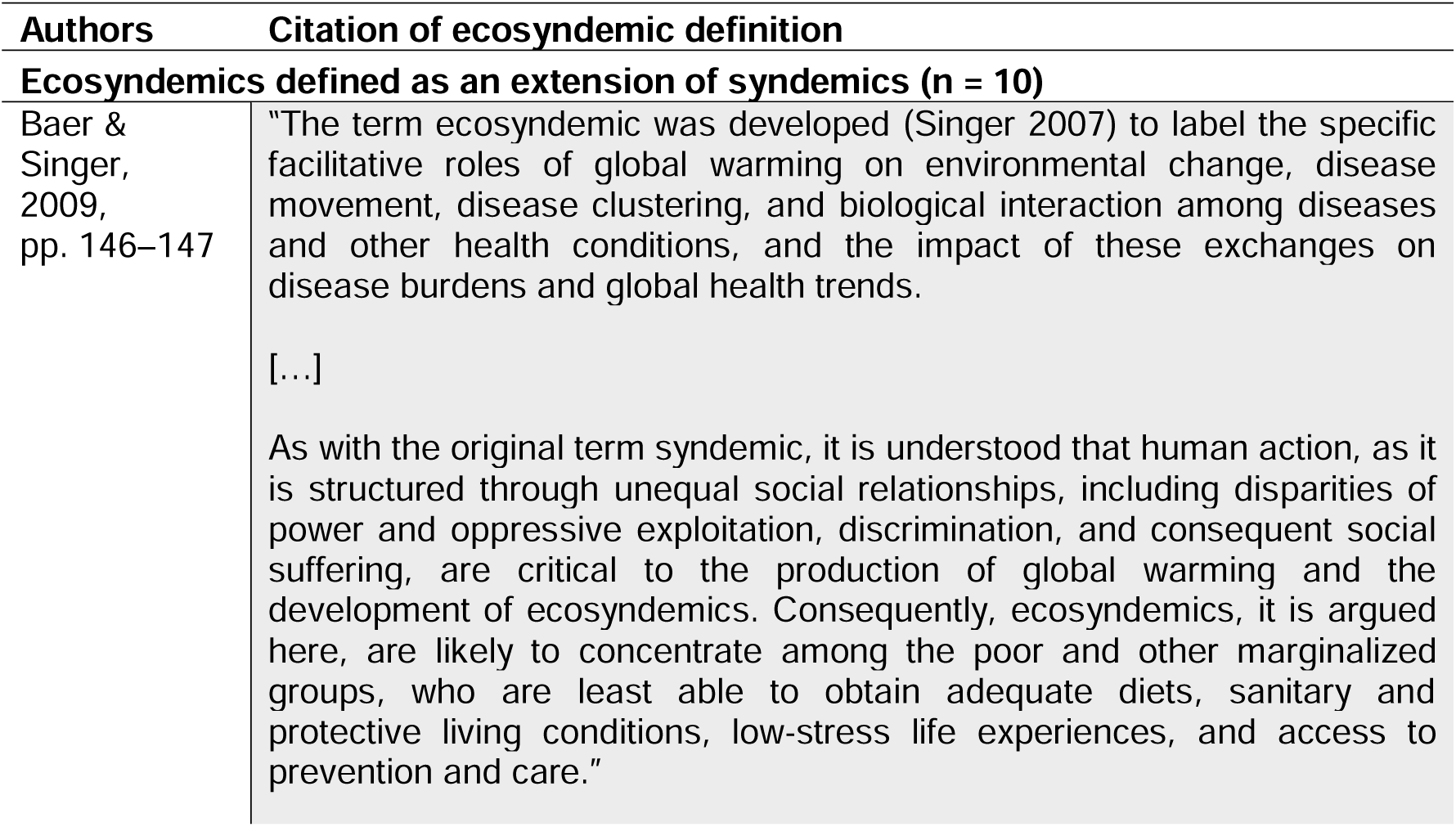
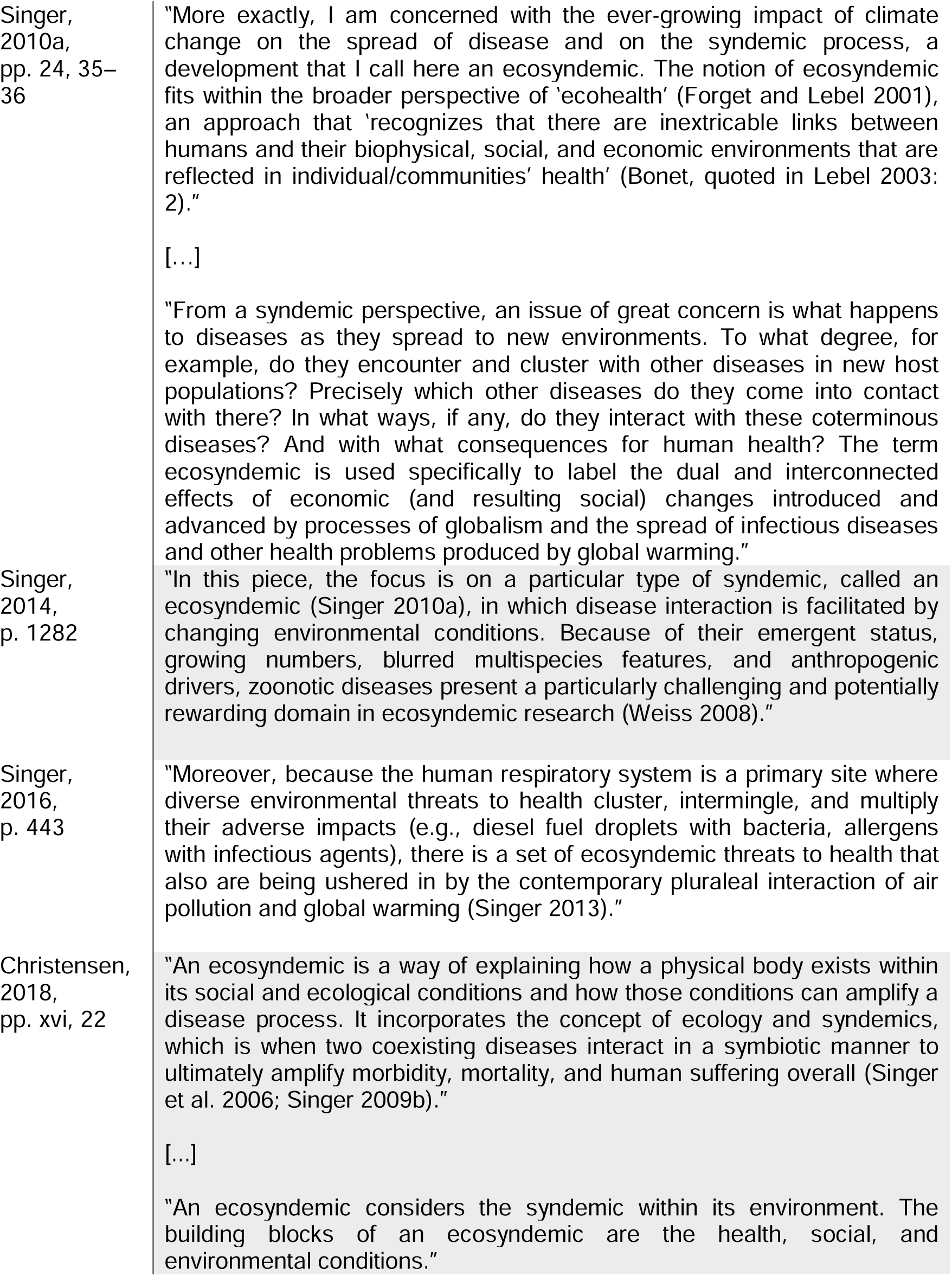
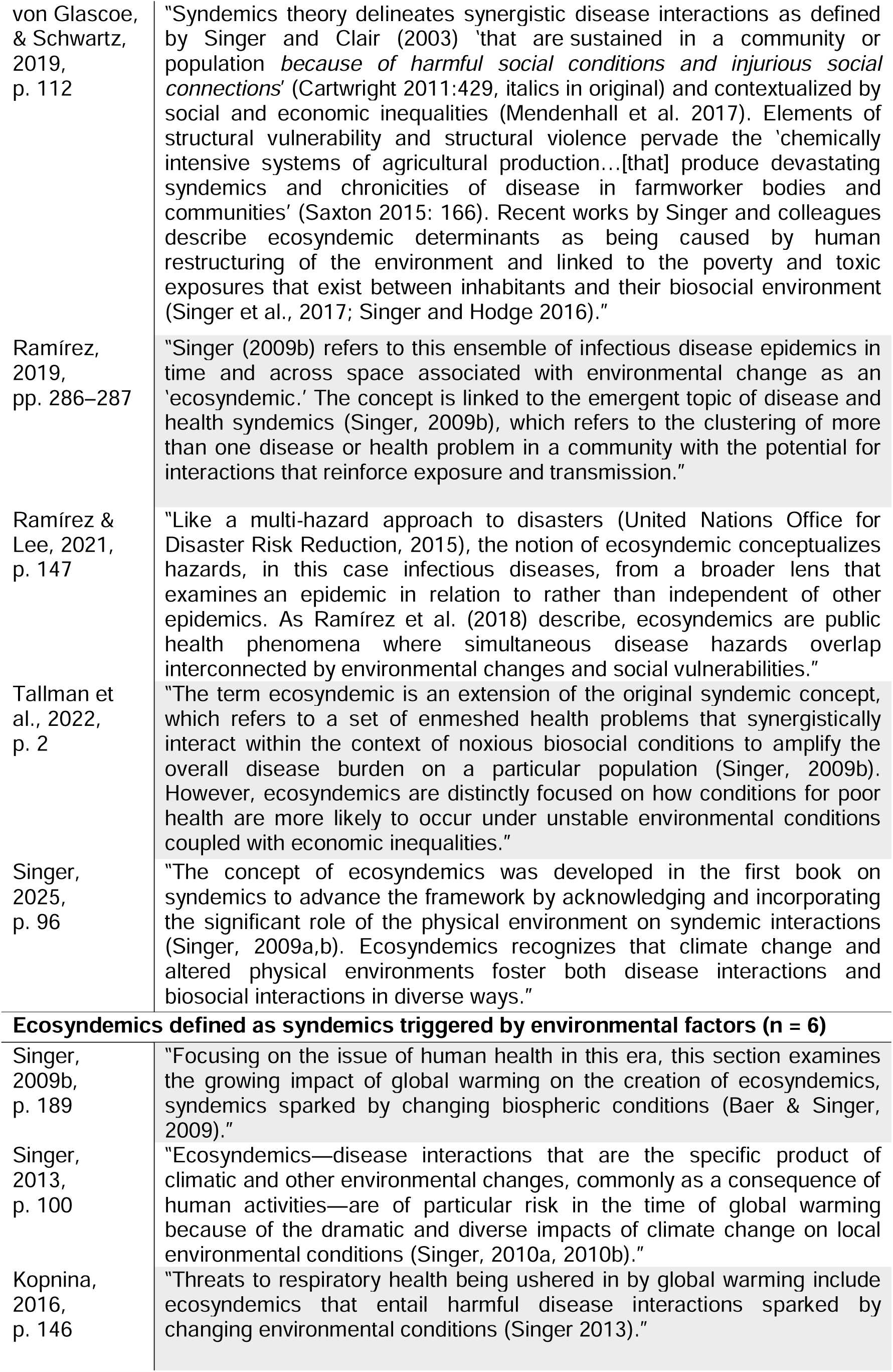
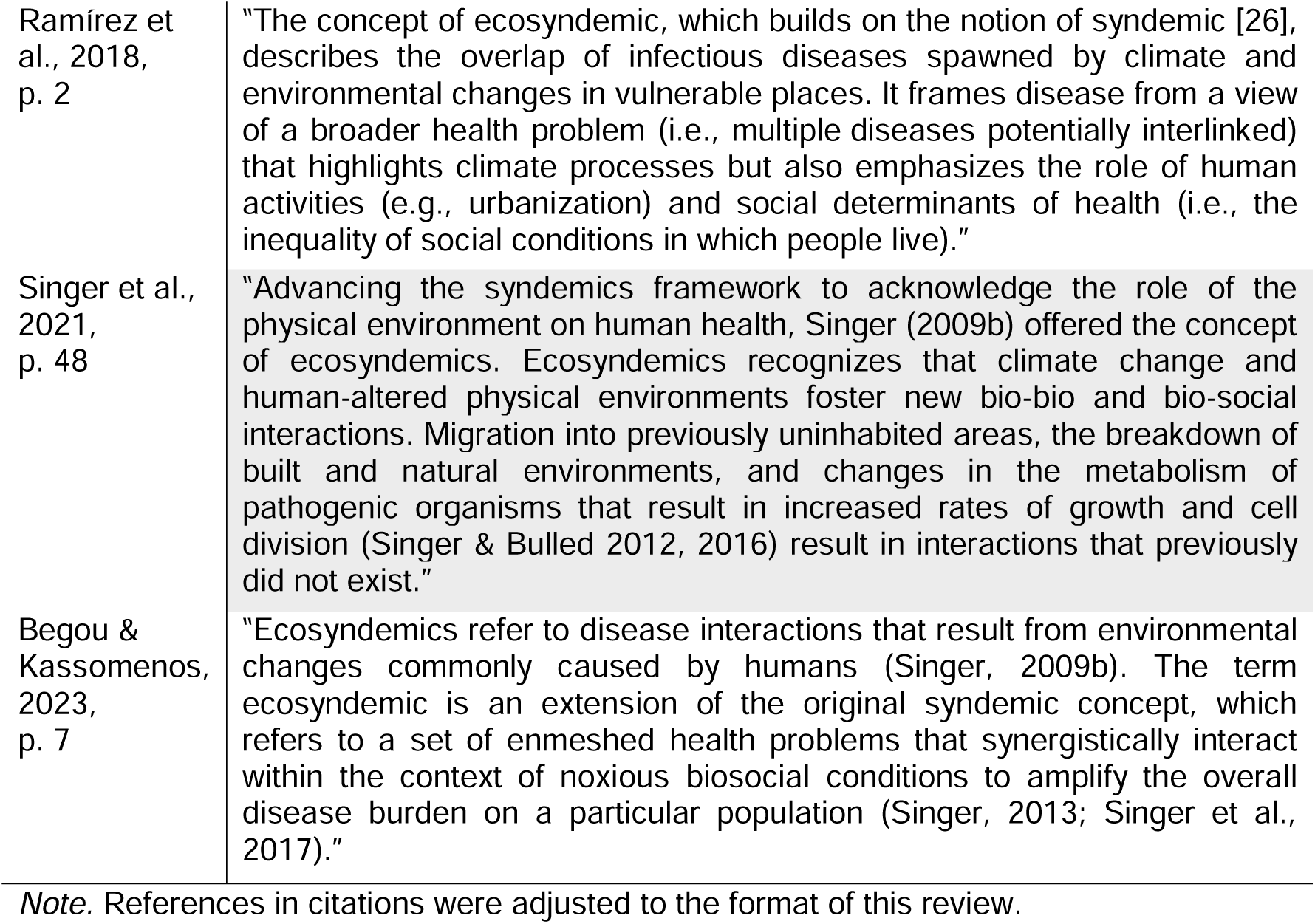
List of Definitions of Ecosyndemics as Cited in Ecosyndemic Publications (n = 16)

In studies that included an extension definition, the aggravation of syndemic outcomes by environmental factors is, using the following terms to refer to the influence of environmental factors: “facilitative,” “interconnected by,” “more likely to occur under unstable environmental conditions,” and “associated with.” This emphasized how environmental factors play a continuous role in ecosyndemics. In the extension definitions, aspects of the physical environment are implemented as contextual factors that exacerbate disease clustering and interaction and amplify population-level health burdens while interacting with social contextual factors.

In publications that included a trigger definition, the temporal aspect of environmental change is emphasized as a trigger after which a syndemic unfolds. The trigger definitions contained terms such as “caused by,” “spawned,” “syndemics that are the product of,” and “sparked by” to emphasize the “shock” effect of environmental change that set an ecosyndemic in motion.

### 4.2 Summary of definitions used

Table 2 presents the code frequencies related to the definitions used within the included empirical and nonempirical studies (n = 169). An ecosyndemic definition was used in 16 publications. The majority of the articles used a syndemic definition (n = 137). The second most frequently used was the global syndemic definition introduced by Swinburn et al. (2019) (n = 16), which centers on the notion that obesity, undernutrition, and climate change interact synergistically on a global scale. A large proportion of the articles were nonempirical (n = 112), in contrast to a smaller proportion of empirical articles (n = 57), five of which used an ecosyndemic definition.

**Table 2.** Code Frequency Table of Ecosyndemic and Syndemic Definitions Used in Ecosyndemic and Syndemic Studies With an Environmental Contextual Focus (n = 169)

|  | **Nonempirical**<br>**(n = 112)** | **Empirical**<br>**(n = 57)** | **Total**<br>**(n = 169)** |
| --- | --- | --- | --- |
| **Definition used** |  |  |  |
| Ecosyndemic | 11 | 5 | 16 |
| Syndemic | 86 | 51 | 137 |
| Global syndemic | 15 | 1 | 16 |
| **Clustering** |  |  |  |
| General clustering | 62 | 36 | 98 |
| Epidemic clustering | 17 | 7 | 24 |
| In time and place | 14 | 7 | 21 |
| Population-level clustering | 10 | 1 | 11 |
| Disease + contributor | 5 | 0 | 5 |
| **Interaction** |  |  |  |
| General disease interaction | 68 | 34 | 102 |
| Biological | 37 | 8 | 45 |
| Social | 17 | 5 | 22 |
| Interaction of epidemics | 17 | 5 | 22 |
| Hybrid | 13 | 4 | 17 |
| Psychological | 9 | 1 | 10 |
| Environmental or localized | 8 | 1 | 9 |
| Behavioral | 7 | 2 | 9 |
| **Context<sup>a</sup>** |  |  |  |
| Socioeconomic | 67 | 33 | 100 |
| Sociocultural | 43 | 36 | 79 |
| Geographical | 50 | 27 | 77 |
| Political | 19 | 7 | 26 |
| Epidemiological | 10 | 9 | 19 |
| Legal | 1 | 1 | 2 |
| Ethical | 0 | 0 | 0 |

<sup>a</sup> Based on the CICI framework (Pfadenhauer et al., 2017).

Regarding clustering, the most frequently used description was that of general clustering (n = 98), which included definitions such as “clustering of multiple diseases” and “the convergence of two or more diseases.” Publications that used an epidemic clustering description (n = 24) highlighted the concurrent or consequential state of multiple epidemics. The time and place clustering description highlights how diseases co-occur in time and place and was used by 21 publications. The population-level description of clustering (n = 11) refers specifically to clustering within as a population-level phenomenon. Finally, the disease plus a contributor description was used in five publications. This description portrays clustering as a phenomenon in which a disease clusters with a contributing factor, such as social problems or environmental threats.

Concerning interaction types, most publications referred to the syndemic criterion of disease interaction as a general disease interaction (n = 102), including descriptions such as the “interaction of two or more diseases,” “disease–disease interactions,” and “two or more disease states that adversely interact with each other.” A substantial number of publications referred specifically to biological interactions (n = 45) consisting of interactions regarding biological pathways, such as a compromised immune system, interspecies interactions, and body alterations that promote the progression of another disease. An interaction of epidemic descriptions, such as interacting epidemics and a synergy of epidemics, was used in 22 publications. A social interaction description, such as social interactions at the interpersonal level or sociocultural and social interactions, was included in 22 publications. A hybrid interaction description was used in 17 publications. The hybrid form combines two interaction types to emphasize specific disease interactions; examples are biosocial, biocultural disease, and bio-psychological interactions. A psychological interaction description was used in 10 studies and a behavioral interaction description in nine publications. Finally, an environmental or localized interaction, including environmental aspects such as climate change and spatial factors, was used in nine publications.

The following contextual factors were mentioned in descending order: the most frequently mentioned were socioeconomic (n = 100), including contextual factors such as income, education, employment, and economic climate. This was followed by sociocultural (n = 79), which included contextual factors related to cultural values, lifestyle, racial discrimination, and historical and contemporary social power relations. The third, geographic contextual factors, were mentioned in 77 publications. Geographic contextual factors include contextual factors related to environmental aspects such as air pollution, climate change, and factors related to the physical environment. Next were political factors (n = 26), including contextual factors such as political authority and ideology. This was followed by epidemiological (n = 19), consisting of contextual factors such as demographics, population density, incidence/prevalence and severity of disease, and morbidity and mortality. Legal factors (n = 2), consisting of contextual factors such as specific legislations and guidelines, were included less frequently. Finally, no publications included ethical contextual factors.

### 4.3 Descriptive summary of empirical studies

A total of 57 empirical studies were identified during the literature search. Appendix B presents a table with an overview of the study characteristics and methods used in the empirical articles. The empirical studies were grouped into quantitative (n = 29), qualitative (n = 15), and mixed (n = 13). The studies were mainly conducted in the US (n = 21), followed by Brazil (n = 4) and the UK (n = 4). The majority of the quantitative studies were cross-sectional (n = 29), followed by longitudinal (n = 12) and cross-sectional pooled (n = 3). Of the mixed studies, eight included a cross-sectional quantitative component and three included a longitudinal quantitative component.

Figure 3 depicts the frequency of inclusion of different environmental factors. Environmental factors related to the built environment were included most frequently (n = 48), followed by those related to the ecological environment (n = 24), the physico-chemical environment (n = 21), and the food environment (n = 18). Specifically, for ecosyndemic studies (n = 5), the built environment (n = 5), ecological environment (n = 5), and physico-chemical environment (n = 5) were included at the same frequency and by all studies, followed by the food environment (n = 2).

**Figure 3.**
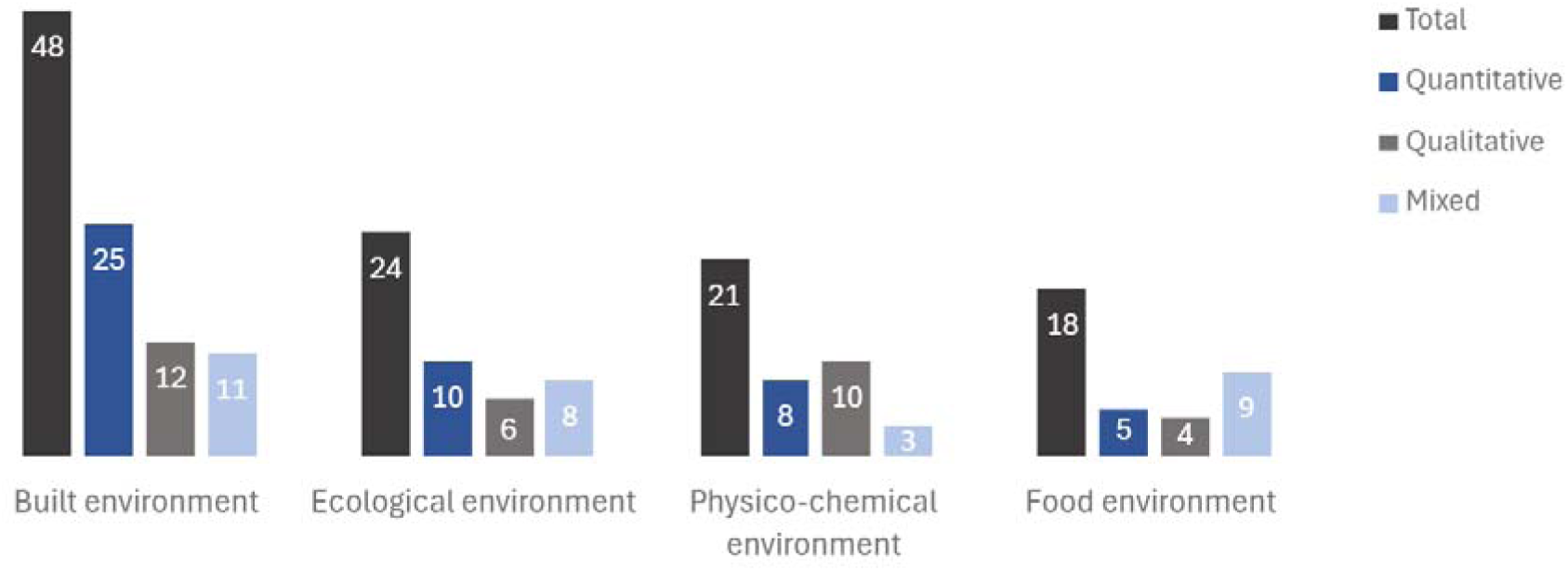
Frequency of Inclusion of Environmental Factor Types in Empirical Studies (n = 57)

### 4.4 Methods used in empirical articles

In Quantitative studies, regression models were used most frequently (n = 25). Environmental factors were included as predictor variables in the regression models. In addition, spatial methods (n = 15) were implemented in several papers to, for example, investigate the association between environmental factors and coinfection incidence or to create spatial visualizations of disease clusters. For both the mixed (n = 13) and qualitative studies (n = 15), the most frequently used methods were interviews (n = 11, n = 11). In interviews, the role of environmental factors was explored by including aspects of the physical environment in the interview guide.

In several papers, distinctive methods were employed to examine syndemics; these included photovoice methods (von Glascoe & Schwartz, 2019), visual ethnography (Tallman et al. 2022), semantic network analysis (Brewis et al. 2022), structured dialogue (Page-Reeves et al. 2019), informal conversations (Slagboom et al. 2022), media content analysis (Mastrangelo et al. 2022), and machine learning (Markovic et al. 2021).

Of the ecosyndemic empirical studies, four used an extension conceptualization, and one employed a trigger conceptualization. For two extension studies, a mixed-methods design was implemented, and for two, a qualitative design was used. The study in which an ecosyndemic trigger framework was implemented used a quantitative design.

**Table 3.** Methods Used in Empirical Articles (n = 57)

|  | **Quantitative**<br>**(n = 29)** | **Mixed**<br>**(n = 13)** | **Qualitative**<br>**(n = 15)** |
| --- | --- | --- | --- |
| Bivariate/multivariate<br>(correlation) | 12 | 7 |  |
| Regression model | 25 | 6 |  |
| Exploratory methods<sup>a</sup><br>(LPA<sup>b</sup>, PCA<sup>c</sup>) | 7 | 0 |  |
| Confirmatory methods<sup>d</sup><br>(SEM<sup>e</sup>, CFA<sup>f</sup>) | 4 | 0 |  |
| Spatial analysis and<br>spatial statistics | 15 | 1 |  |
| Interviews |  | 11 | 11 |
| Focus groups |  | 4 | 3 |
| Participant observation |  | 3 | 3 |
| Ethnographic fieldwork |  | 1 | 3 |
| Historical sources |  | 1 | 1 |
| Other | 1 | 4 | 2 |

<sup>a</sup> Methods used to explore underlying relationships in data.<sup>b</sup> Latent profile analysis.<sup>c</sup> Principal component analysis.<sup>d</sup> Methods used to confirm potential hypotheses.<sup>e</sup> Structural equation model.<sup>f</sup> Confirmatory factor analysis.**Table 4***Methods Used in Ecosyndemic Empirical Articles (n = 5)*

**Table 4.** Methods Used in Ecosyndemic Empirical Articles (n = 5)

| **Ecosyndemics defined as an extension of syndemics (n = 4)** |  |
| --- | --- |
| Christensen, 2018 | Interviews, participant observation, ethnographic fieldwork |
| Von Glascoe et al.,<br>2019 | Interviews, participant observation, photovoice |
| Ramírez, 2019 | GIS<sup>a</sup>, descriptive data |
| Tallman et al.,<br>2022 | Key-informant interviews, bivariate analysis, focus groups,<br>visual ethnography, document collection |
| **Ecosyndemics defined as syndemics triggered by environmental factors (n = 1)** |  |
| Ramírez et al.,<br>2018 | Geographic information system for spatial visualizations,<br>Global Moran's I, Getis-Ord Gi hotspot analysis, PCA<sup>b</sup>,<br>Pearson's correlation |

<sup>a</sup> Geographic information system.<sup>b</sup> Principal component analysis.

## 5. Discussion

This scoping review examined how syndemic research engaged with environmental contextual factors. Specifically, the study explored how ecosyndemics are defined and how environmental contextual factors are studied, operationalized, and conceptualized. We identified two conceptualizations of ecosyndemics: the *extension* conceptualization, used across both ecosyndemic and syndemic studies, and the *trigger* conceptualization, exclusively used in ecosyndemic studies.

Before interpreting our findings in light of the broader literature, we reflect on the observation that most of the studies included in this review were nonempirical. Of all the publications reviewed, only five were empirical ecosyndemic studies, suggesting that ecosyndemic research remains primarily conceptual.

One possible explanation for the high number of nonempirical publications is the origin of syndemics theory, which is a critical theory that is rooted in medical anthropology and that questions the biomedical paradigm that focuses on individual health conditions separately. We observed that nonempirical studies often used a syndemic lens to increase understanding or awareness of the link between environmental factors and clustering and the adverse interactions of diseases or to increase awareness of syndemic vulnerability from a biosocial complex perspective. Another possible explanation lies in the increased adoption of syndemics theory: The research field has grown immensely since its introduction, thereby shifting toward integrating different scientific epistemologies, leading to methodological heterogeneity (Pirrone et al., 2021; Gizamba et al., 2023). In light of the adoption of the ecosyndemics concept into different research fields, operationalizing ecosyndemic theory into empirical research might prove to be a complex process for researchers. Struggles over the operationalization of contextual factors in syndemic studies have been well documented in recent research (Pirrone et al., 2021; Gizamba et al., 2023).

The operationalization of contextual factors comes with added complexity for ecosyndemics, particularly when both macro-scale environmental and social factors are involved. A possible solution lies in understanding how ecosyndemics are conceptualized in current research, such as how, for example, the identification of the two ecosyndemic concepts aid in understanding how ecosyndemics differ from syndemics with an environmental contextual focus. The *extension* concept emphasizes the role of the environmental context in combination with other contextual factors, such as poverty, stigma, and structural violence. However, the extension conceptualization is used in both ecosyndemic and syndemic studies with an environmental contextual focus, as certain syndemic studies with this focus apply the ecosyndemic extension conceptualization without using the specific term. In contrast, the *trigger* conceptualization is used solely within ecosyndemic studies because the environmental context is conceptualized differently from the traditional syndemic framework—namely, as a trigger to a syndemic. Regarding the operationalization of contextual factors, the trigger conceptualization differs from the extension conceptualization from a temporal perspective. In the trigger conceptualization, a syndemic can be set in motion in two ways. In the first, a buildup of environmental exposures, such as exposure to pesticides (von Glascoe & Schwartz, 2019), accumulate into a tipping point which, when surpasses, sets a syndemic in motion. In the second, a sudden event such as extreme precipitation (Ramírez et al., 2018) acts as trigger to the onset of a syndemic.

The trigger conceptualization enables the analysis of environmental factors from two perspectives: how environmental factors trigger an ecosyndemic and how the environmental context changes once an ecosyndemic has been triggered. The study by Tallman et al. (2022) is an example of how taking the trigger as a lens can aid in the operationalization of contextual factors. The study uses an extension definition, but the health effects of large construction projects that are the topic of study correspond with the trigger conceptualization in which the construction of a dam and highway act as a trigger for a plethora of negative health outcomes for the local populations. However, the issue here is that the construction projects were a sudden event and not an accumulation of environmental exposures, such as that which occurs in trigger definitions that focus on the effects of global warming on ecosyndemics (Singer, 2009b). Further distinctions thus need to be made to pinpoint the exact position of not only ecosyndemics in the field but also of the two separate conceptualizations.

The methods used in ecosyndemic and syndemic studies also highlight how the operationalization of contextual factors is divided into different scales: micro, meso, and macro. An indication of this is that regression models were included most frequently (n = 31) among the empirical studies that were included in this review. A possible explanation for this is that regression analysis is suited for the operationalization of environmental factors at the micro level, such as residential classification (urban/suburban/rural), population density, supermarket density, whose operationalization is less complex than that of meso and macro factors. Supporting this argument is the finding that studies that focus on macro-level factors, such as the effects of laws and policies on working conditions and work-related health problems, are qualitative in nature. For example, Horton (2016) focused on the effects of public policies on farmworkers’ health in relation to heat stroke, and Unterberger (2018) studied how lack of workplace safety and health regulations lead to work-related injuries among Latino immigrant workers in the USA. For meso-scale factors, this distinction is less evident, as they were incorporated into the analysis of both quantitative and qualitative studies. For example, Frye et al.’s (2014) qualitative study examined how neighborhood space affects social disparities in sexual health outcomes, and a quantitative study by Galeana-Pizaña et al. (2023) focused on how neighborhood food environments are related to mortality from diseases associated with poor nutrition. The following question thus arises: To what extent should a syndemic study include contextual factors for all three scales— micro, meso, and macro? Ecosyndemic and syndemic studies that use a quantitative design might encounter issues with the operationalization of macro-scale factors and are less suited for capturing human interactions and experiences. In contrast, studies that employ a qualitative design are less equipped to address questions regarding correlations and population-level health outcomes. A mixed-methods design can be a possible bridge between these approaches. However, the complexity of syndemics makes it difficult to grasp contextual factors on different scales within a single study while also fulfilling the first and second syndemic criteria through the inclusion of a detailed analysis of population-level disease clustering and interaction. Although mixed-methods studies can partially provide insight into clustering, interaction, and context, there is a trade-off in which the wider scope of methods used comes at a cost. The mixed-methods studies included in this review, for example, did not use advanced statistical methods such as PCA and SEM. Therefore, an investigation into a possible syndemic might require multiple studies in which clustering and interaction are investigated and then followed up by in-depth qualitative research to uncover the underlying contextual patterns that drive the syndemic.

Although the environmental focus of ecosyndemics aids in highlighting the role of environmental factors in relation to disease clustering and interaction, the social context remains an essential component of ecosyndemics. The code frequency analysis shows that geographic contextual factors are included third most frequently in ecosyndemic and syndemic definitions. However, the most frequently mentioned contextual factors were related to the socioeconomic context, which is in line with the findings of reviews by Gizamba et al. (2023) and Pirrone et al. (2021). One potential reason for the higher inclusion of socioeconomic and sociocultural factors within syndemic definitions is that even in ecosyndemics, the geographic context is always an added layer to the core syndemic contextual considerations that are related to socioeconomic and sociocultural contextual aspects. Socioeconomic factors are emphasized within ecosyndemic definitions that often focus on the combined effect of environmental and socioeconomic contextual aspects, such as poverty and vulnerability. Although ecosyndemics as a separate definition from syndemics highlights the effects of environmental factors on the latter, we believe that the underlying social context must always remain part of this focus because the interaction between the environmental and social contexts are the key component to understanding the population-level health outcomes and health inequalities caused by ecosyndemics.

## 6. Strengths and limitations

One strength of this review is the interdisciplinary approach that was taken regarding the inclusion and analysis of publications. This allowed us to examine the use of ecosyndemics and syndemics within different scientific disciplines. An additional strength is that we used a broad strategy to conduct searches across multiple databases to include a wide range of syndemic publications with an environmental contextual focus. The inclusion of a high number of publications enabled an understanding of the multifaceted aspects of ecosyndemics and syndemics.

This review has several limitations. First, the relatively low number of empirical ecosyndemic studies (n = 5) makes the interpretation of the review findings complex, thereby complicating any conclusive answer regarding the operationalization of contextual factors and methodology. Second, categorizing contextual factors through the CICI framework (Pfaddenhauer et al., 2017) and environmental factors through the exposome framework (Beulens et al., 2022) might have led to a more funneled categorization of contextual and environmental factors. For environmental factors in particular, the four types included in this review—the built environment, ecological environment, physico-chemical environment, and food environment—might not cover all aspects of the physical environment. Moreover, some categories, such as the built environment, encompass larger areas of the physical environmental domain compared to other categories, such as the food environment, which might have led to an overcategorization of environmental factors in the built environment category. Therefore, the interpretation of the results regarding environmental factor categories should take into account the varying breadths of the categories.

## 7. Future outlooks

Various methods were used to investigate syndemics in the studies that were included in this review. To grasp the complexity of ecosyndemics, novel frameworks and methods could be implemented in future research. One way to account for the complexity of contextual factors interacting with one another, as well as with health conditions, is through a systems thinking approach. The aim of systems thinking is to understand how elements are connected to one another as part of a larger whole (Peters, 2014). Therefore, it can arguably fit a syndemic scope in which diseases and contextual factors continuously interact with one another. Systems thinking includes a range of methods for mapping connections between elements. These methods include causal loop diagrams, system dynamics, and agent-based modeling. Systems thinking might also offer opportunities regarding operationalization by mapping the interactions between factors instead of regarding them as individual elements for analysis. Through a systems approach, researchers can identify and study factors that initially do not have an obvious influence on syndemic interactions and health outcomes. For example, Deutsch et al. (2022) used system dynamics to include personal experiences and to examine syndemic interactions regarding stigmatized public health issues. The final model included factors on the micro, meso, and macro scales, showing potential new possibilities for syndemic research based on systems thinking.

## 8. Conclusion

The concept of ecosyndemics can emphasize the role of the physical environment in addition to the classical conceptualization of the syndemics of disease clustering and disease interaction in adverse social contexts. It can also support the discussion by showing how health and the environment are interconnected and should not be seen as separate from each other. The ecosyndemic framework can offer insight into policy recommendations that focus on the prevention of unequal population-level health burdens through interventions that focus on adverse living environments and social contexts. Most studies included in this review were nonempirical in nature. This might be related to issues regarding the operationalization of contextual factors and the complexity of studying multiple such factors and interactions being barriers to conducting empirical research on ecosyndemics. Because the study of ecosyndemics requires a multidisciplinary approach consisting of various methods, it is essential, where possible, for the triangulation of the results achieved through various methods to be included within ecosyndemic studies. There is much potential for ecosyndemic empirical research, as it is an understudied field that can provide insight into the complex interactions between the physical and social domains with regard to population-level disease clustering.

## Supporting information

Supplemental references empirical articles

Supplemental references nonempirical articles

## Data Availability

All data produced in the present work are contained in the manuscript.

## Acknowledgments

In memoriam to Merrill Singer. We thank Professor Singer for his thoughtful insights during the first phases of this project.

We would like to acknowledge the Walaeus library team and in particular C.F. Pees for her help in drafting the search queries used in this review.

## Appendix

## Appendix A Search Queries Used for the Consulted Databases

### Query strings

An experienced information expert from the Leiden University Medical Centre (LUMC) was consulted in order to help create a search strategy. The search strategy was created iteratively by using preliminary searches and comparing result from different search queries. This resulted in the following final search terms:

**PubMed**: “ecosyndemic*”[All Fields] OR ((“syndemic*”[MeSH Terms] OR “syndemic*”[All Fields]) AND (“Ecology”[MeSH Terms] OR “Environment”[MeSH Terms] OR “Industry”[MeSH Terms] OR “agriculture*”[MeSH Terms] OR “Housing”[MeSH Terms] OR “Social Environment”[MeSH Terms] OR “eco”[All Fields] OR “ecolog*”[All Fields] OR “anthrop*”[All Fields] OR “environ”[All Fields] OR “environs”[All Fields] OR “environment*”[All Fields] OR “urban*”[All Fields] OR “neighborhood*”[All Fields] OR “neighbourhood*”[All Fields] OR “communit*”[All Fields] OR “city”[All Fields] OR “cities”[All Fields] OR “rural*”[All Fields] OR “spatial*”[All Fields]))

**Embase**: “ecosyndemic*”.mp. OR ((exp “syndemic”/ OR “syndemic*”.mp.) AND (exp “Ecology”/ OR exp “Environment”/ OR exp “Industry”/ OR exp “agriculture”/ OR exp “Housing”/ OR exp “Social Environment”/ OR “eco”.mp. OR “ecolog*”.mp. OR “anthrop*”.mp. OR “environ”.mp. OR “environs”.mp. OR “environment*”.mp. OR “urban*”.mp. OR “neighborhood*”.mp. OR “neighbourhood*”.mp. OR “communit*”.mp. OR “city”.mp. OR “cities”.mp. OR “rural*”.mp. OR “spatial*”.mp.))

**ScienceDirect**: The interface used by ScienceDirect does not allow more than 8 Boolean operators or the use of a ‘*’ wildcard. To obtain results that are comparable to the four separate searches in the other four databases, four separate searches were conducted with different search queries.

1. ”ecosyndemic” OR “ecosyndemics” OR ((“syndemic” OR “syndemics”) AND (“Industry” OR “agriculture” OR “eco” OR “environmental” OR “community”))
2. ”ecosyndemic” OR “ecosyndemics” OR ((“syndemic” OR “syndemics”) AND (“Ecology” OR “Housing” OR “Social Environment” OR “ anthropocene” OR “urban”))
3. ”ecosyndemic” OR “ecosyndemics” OR ((“syndemic” OR “syndemics”) AND (“neighborhood” OR “neighbourhood” OR “city” OR “cities” OR “rural”))
4. ”ecosyndemic” OR “ecosyndemics” OR ((“syndemic” OR “syndemics”) AND (“spatial”))

**Web of Science**: TS=”ecosyndemic*” OR (TS=(“syndemic” OR “syndemic*”) AND TS=(“Ecology” OR “Environment” OR “Industry” OR “agriculture” OR “Housing” OR “Social Environment” OR “eco” OR “ecolog*” OR “anthrop*” OR “environ” OR “environs” OR “environment*” OR “urban*” OR “neighborhood*” OR “neighbourhood*” OR “communit*” OR “city” OR “cities” OR “rural*” OR “spatial*”))

**PsycINFO**: TX “ecosyndemic*” OR ((TX “syndemic*”) AND (DE “Ecology” OR DE “Behavioral Ecology” OR DE “Ecological Factors” OR DE “Social Ecology” OR DE “Environment” OR DE “Built Environment” OR DE “Environmental Enrichment” OR DE “Environmental Planning” OR DE “Facility Environment” OR DE “Learning Environment” OR DE “Nature (Environment)” OR DE “Person Environment Fit” OR DE “Physical Comfort” OR DE “Public Space” OR DE “Single Sex Environments” OR DE “Social Environments” OR DE “Sustainability” OR DE “Therapeutic Environment” OR DE “Virtual Environment” OR DE “Industrialization” OR DE “Industrial Accidents” OR DE “Agriculture” OR DE “Housing” OR DE “Assisted Living” OR DE “Dormitories” OR DE “Group Homes” OR DE “Retirement Communities” OR DE “Shelters” OR DE “Social Environments” OR DE “Academic Environment” OR DE “Animal Environments” OR DE “Communities” OR DE “Home Environment” OR DE “Poverty Areas” OR DE “Rural Environments” OR DE “Suburban Environments” OR DE “Towns” OR DE “Urban Environments” OR DE “Working Conditions” OR DE “Environmental Stress” OR DE “Nature Nurture” OR DE “Urban Planning” OR TX “eco” OR TX “ecolog*” OR TX “anthrop*” OR TX “environ” OR TX “environs” OR TX “environment*” OR TX “urban*” OR TX “neighborhood*” OR TX “neighbourhood*” OR TX “communit*” OR TX “city” OR TX “cities” OR TX “rural*” OR TX “spatial*”))

## Appendix B

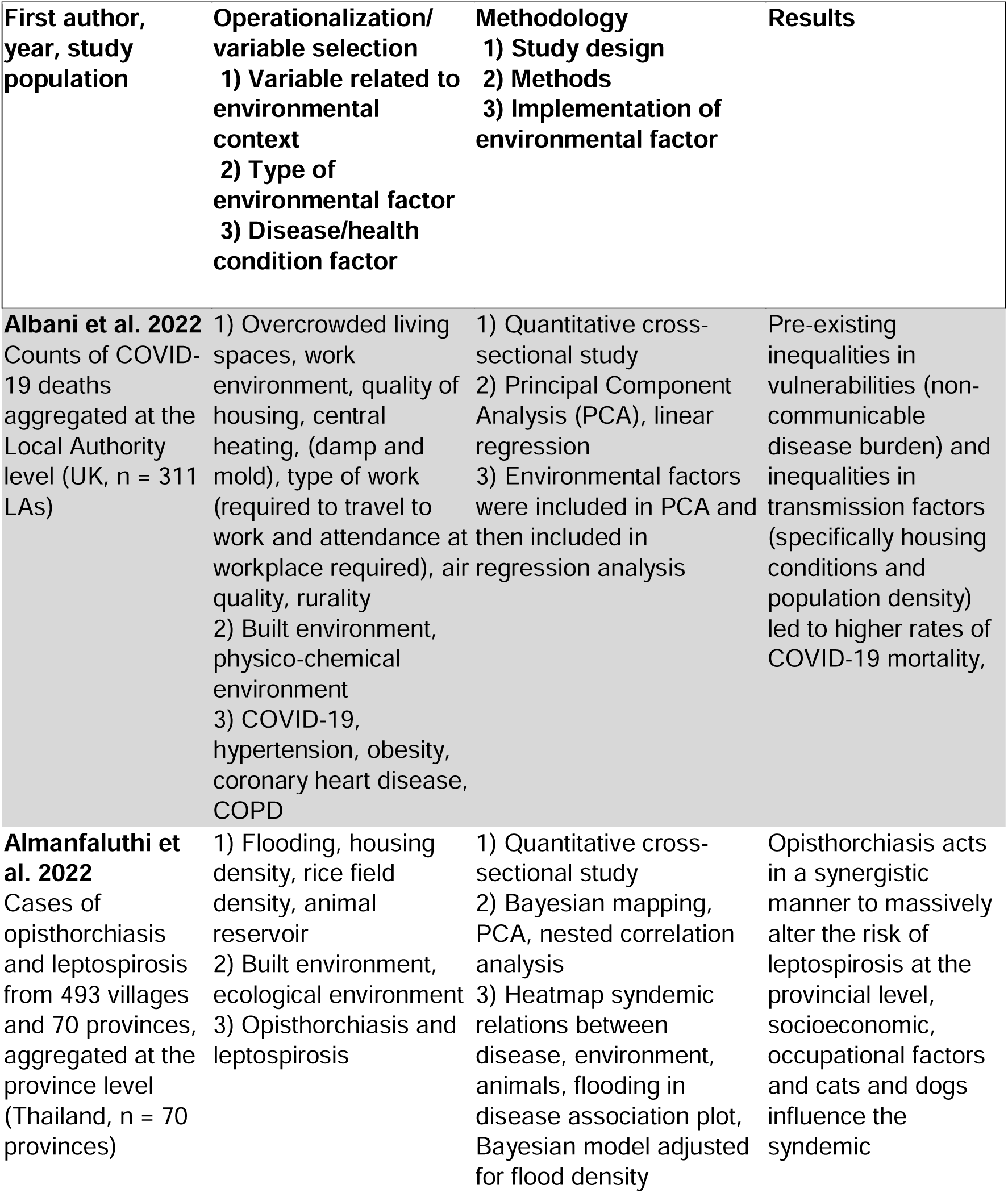
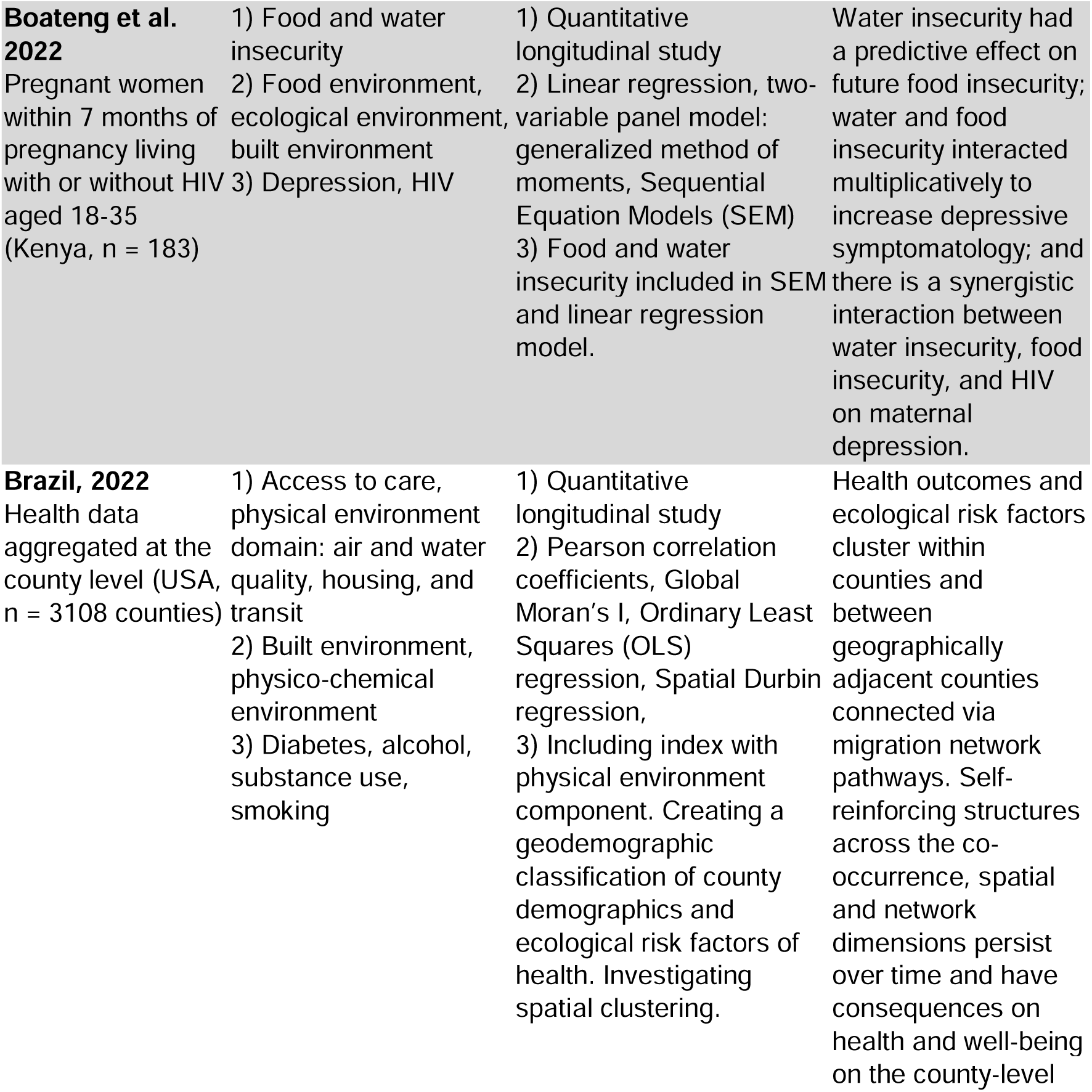
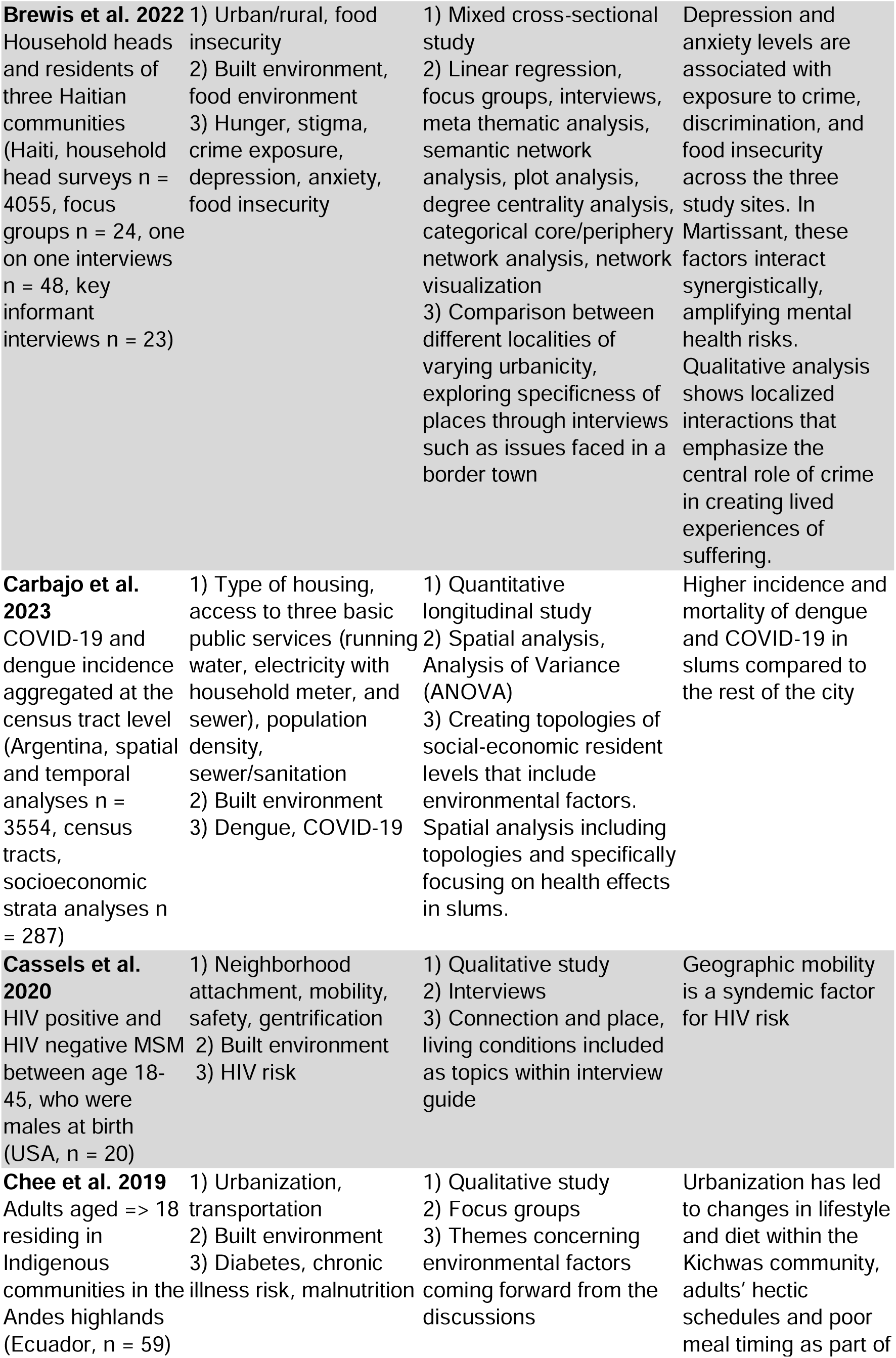
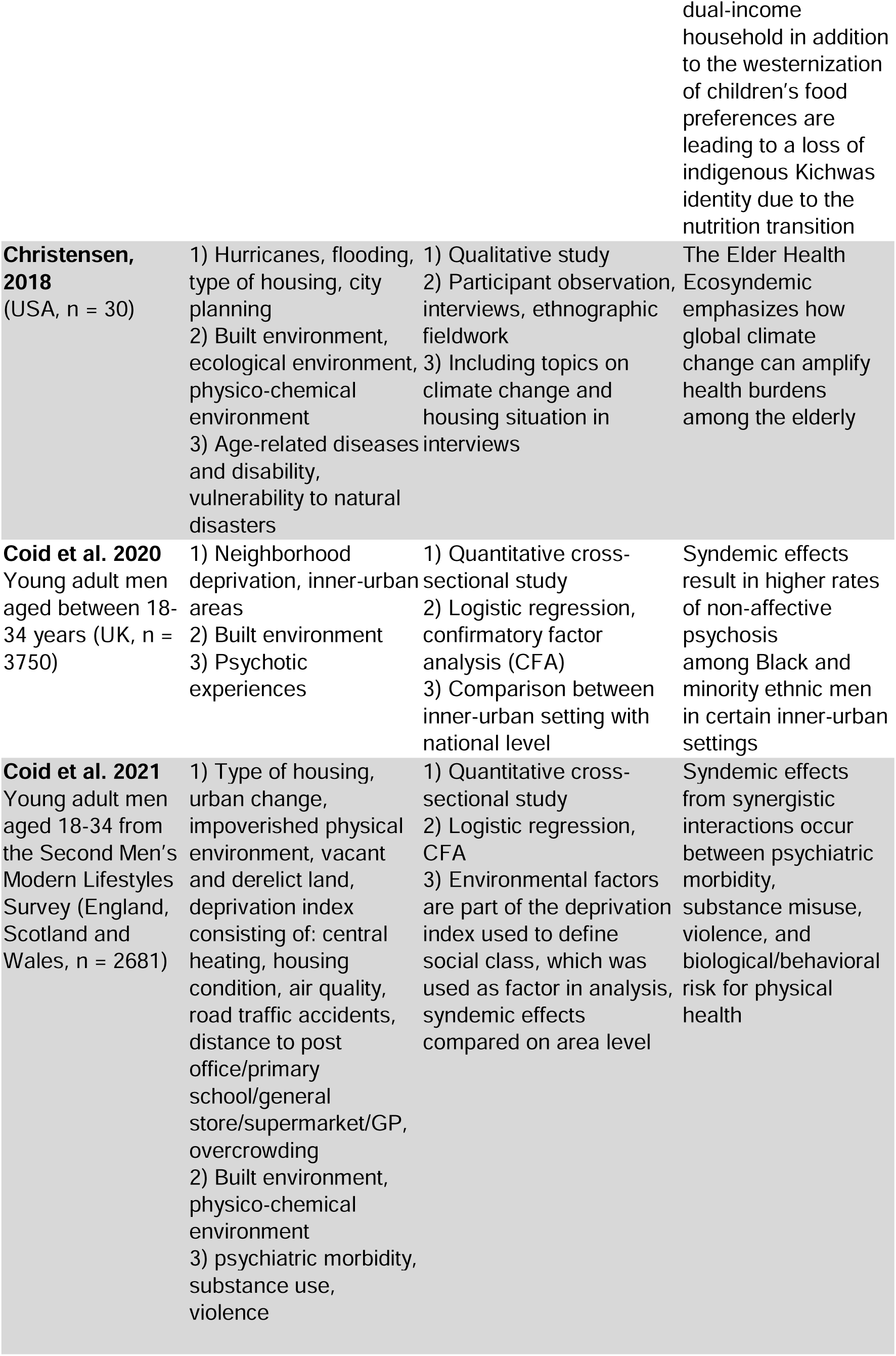
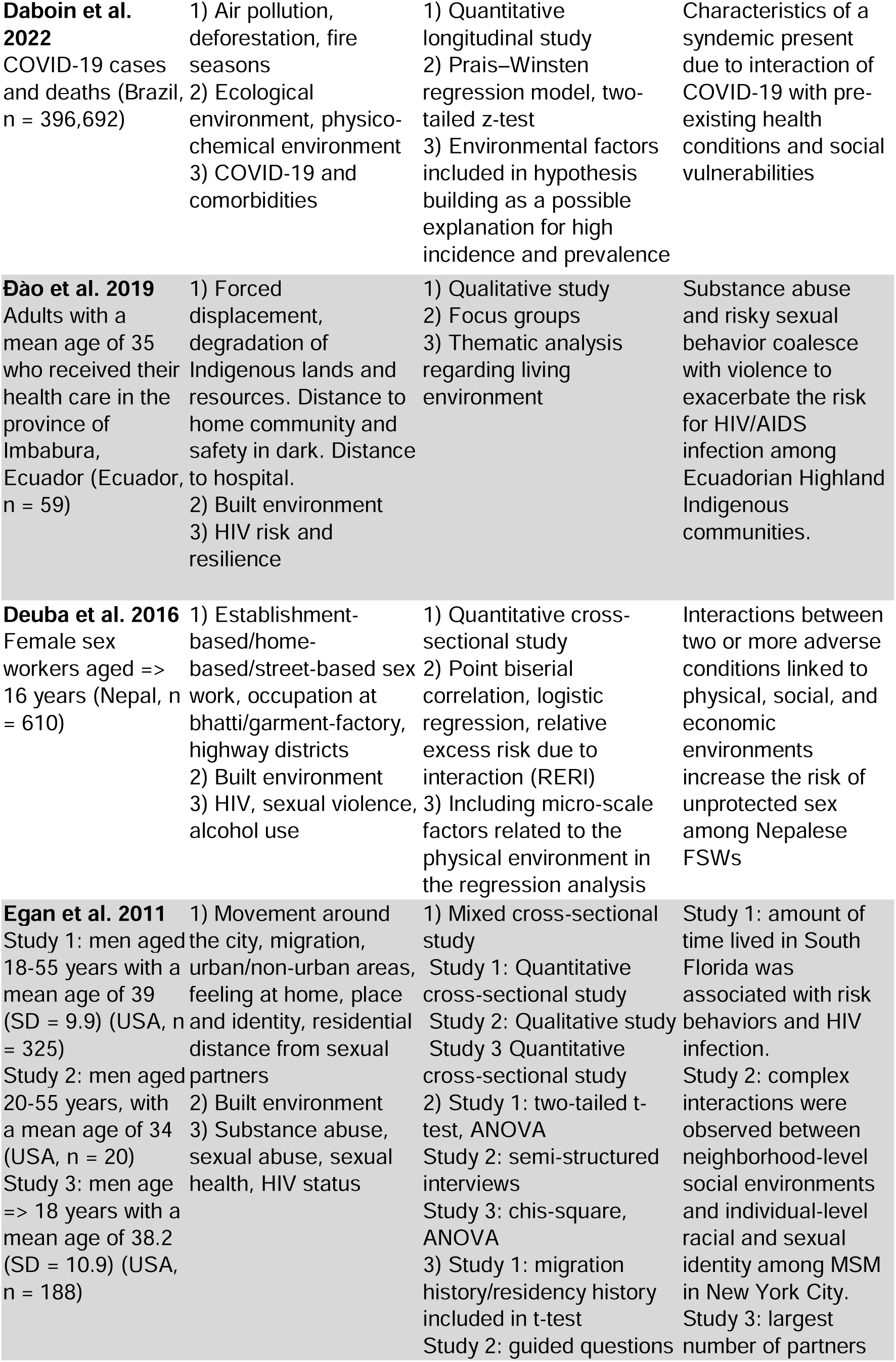
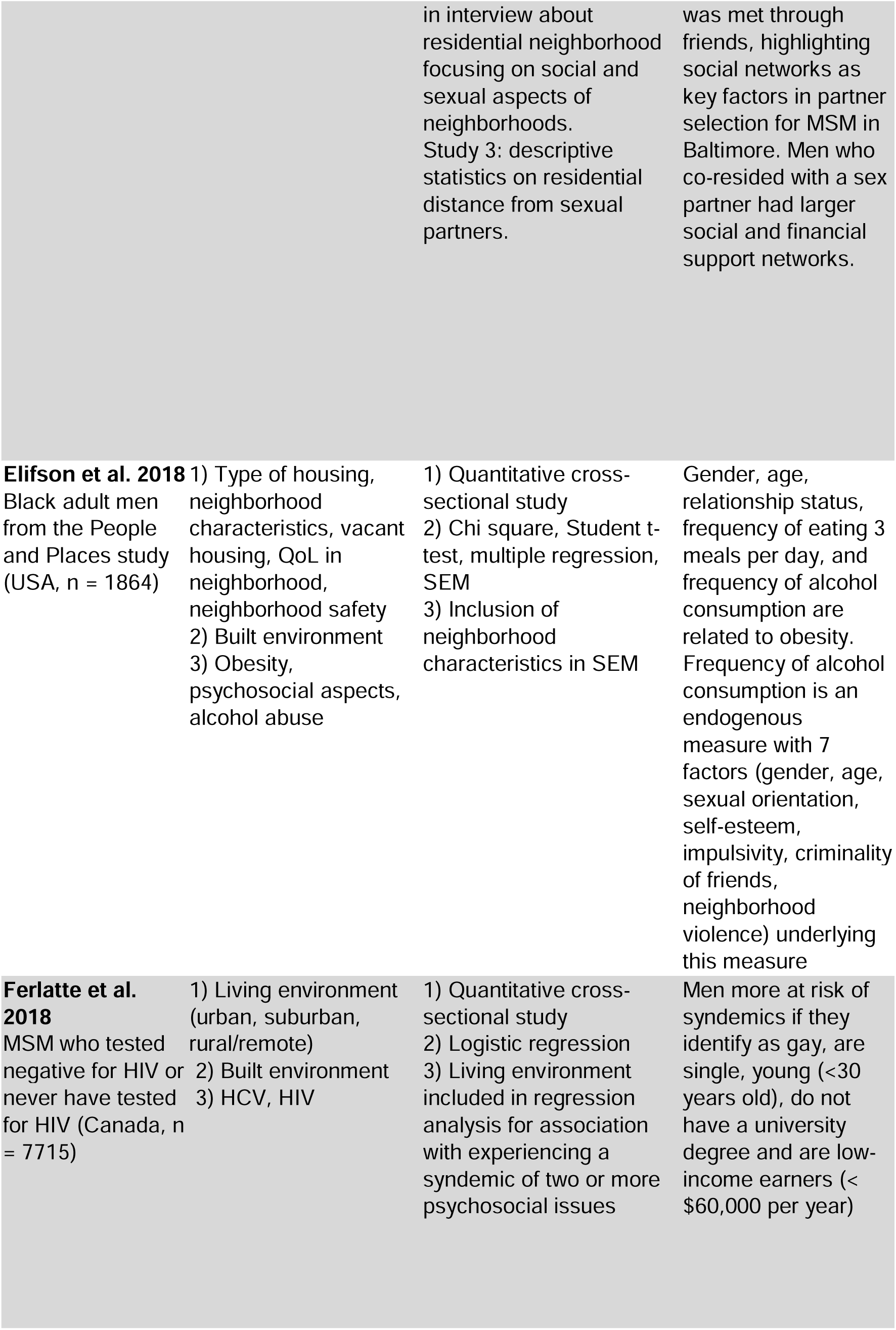
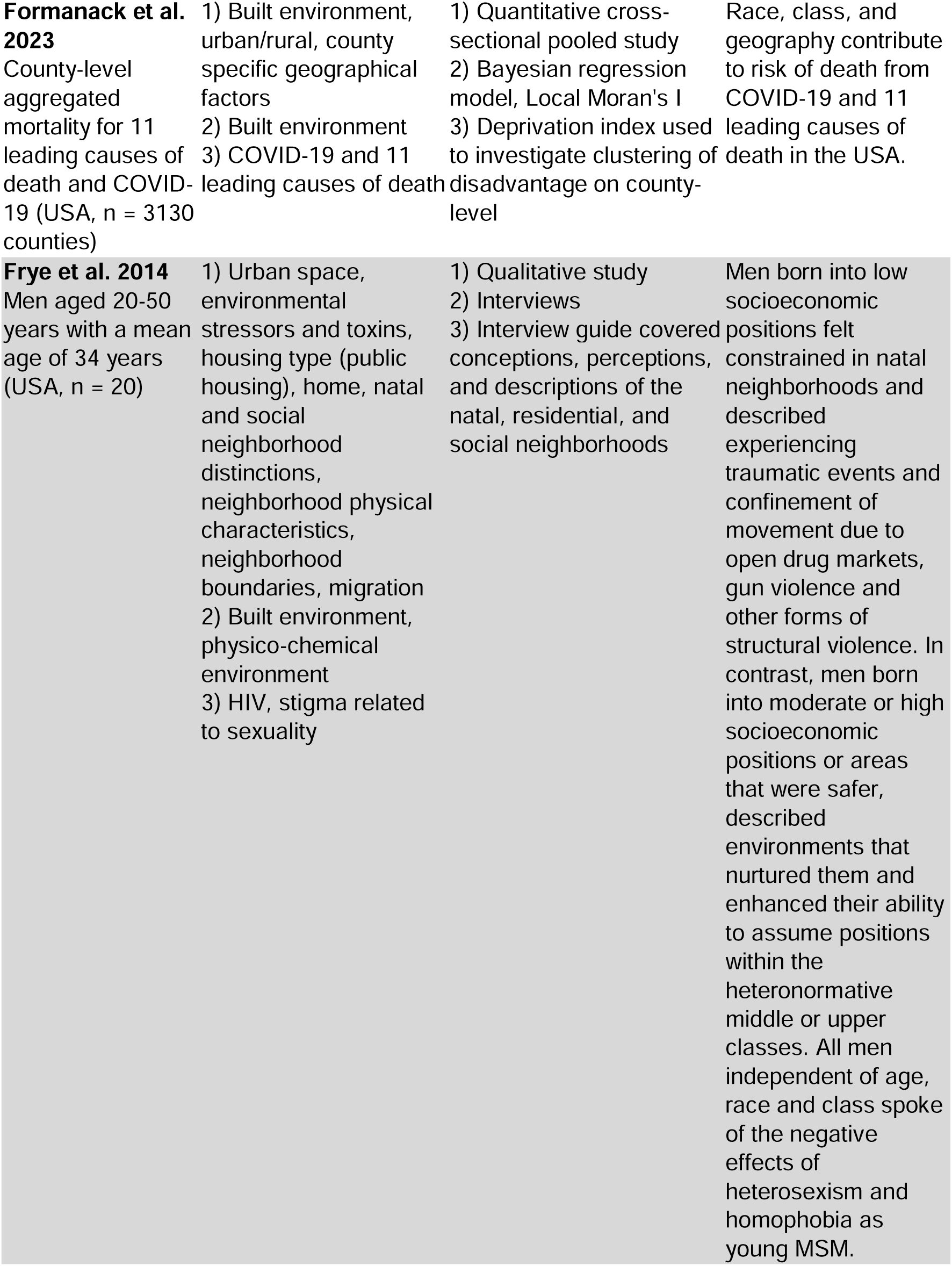
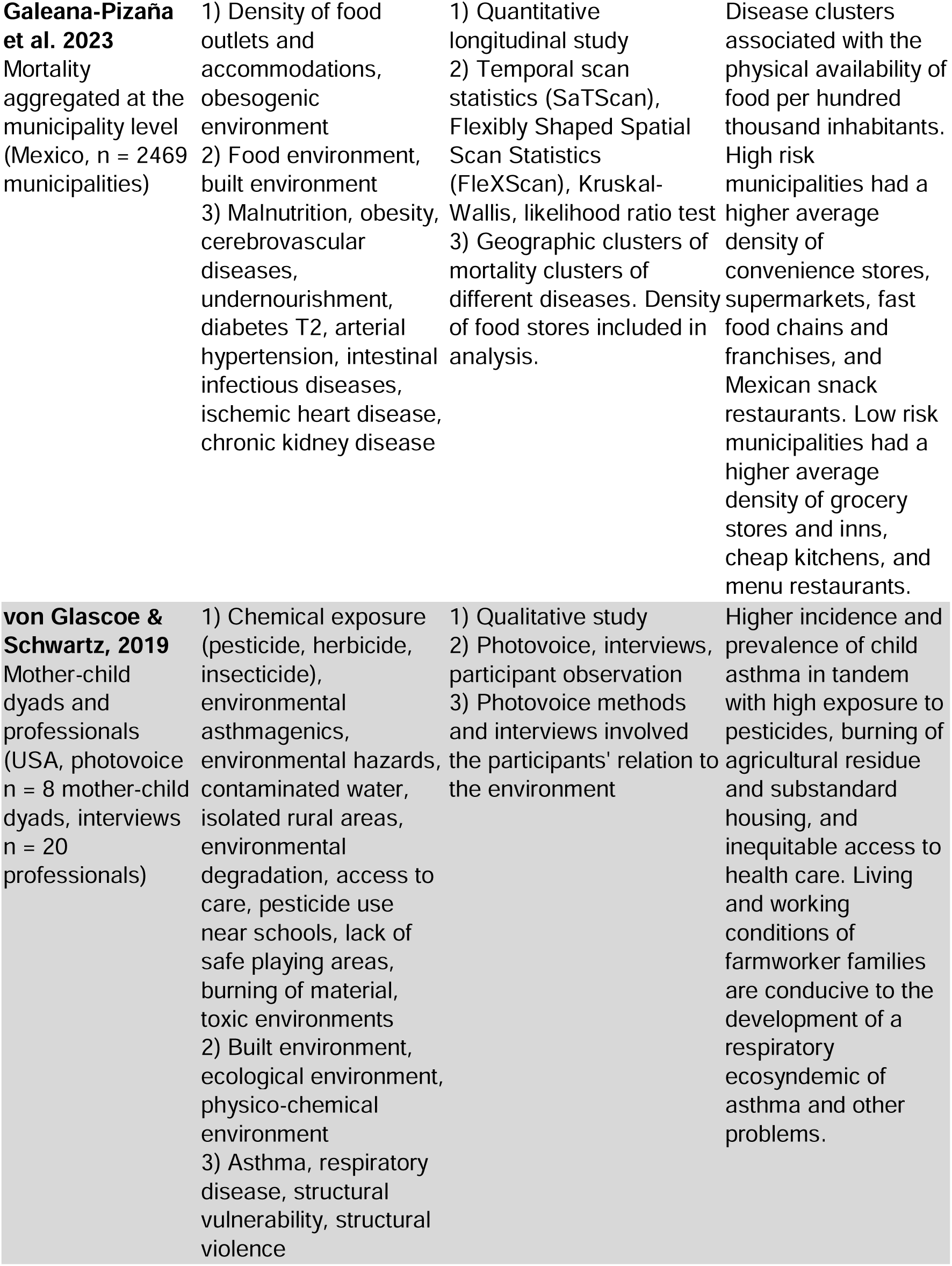
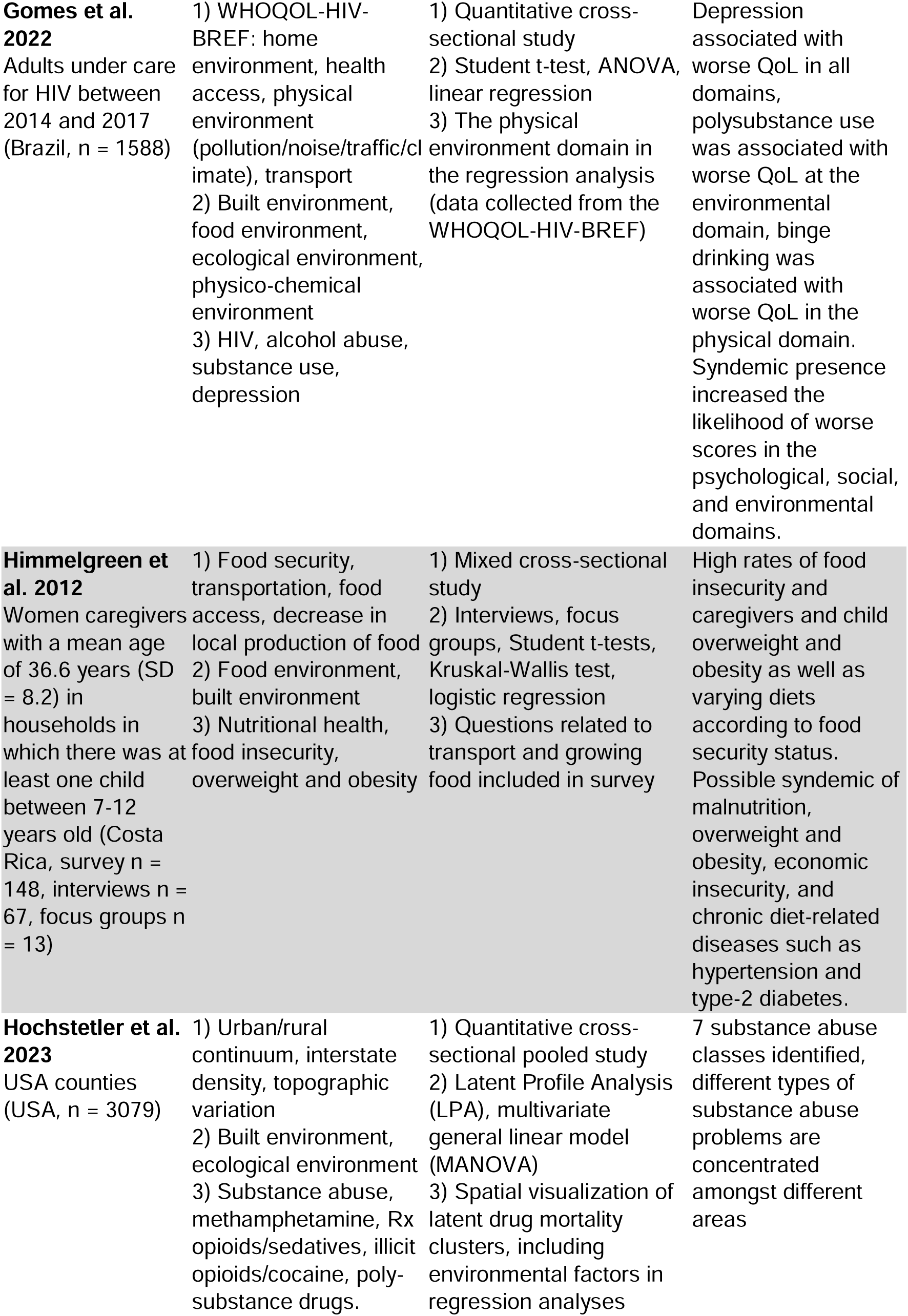
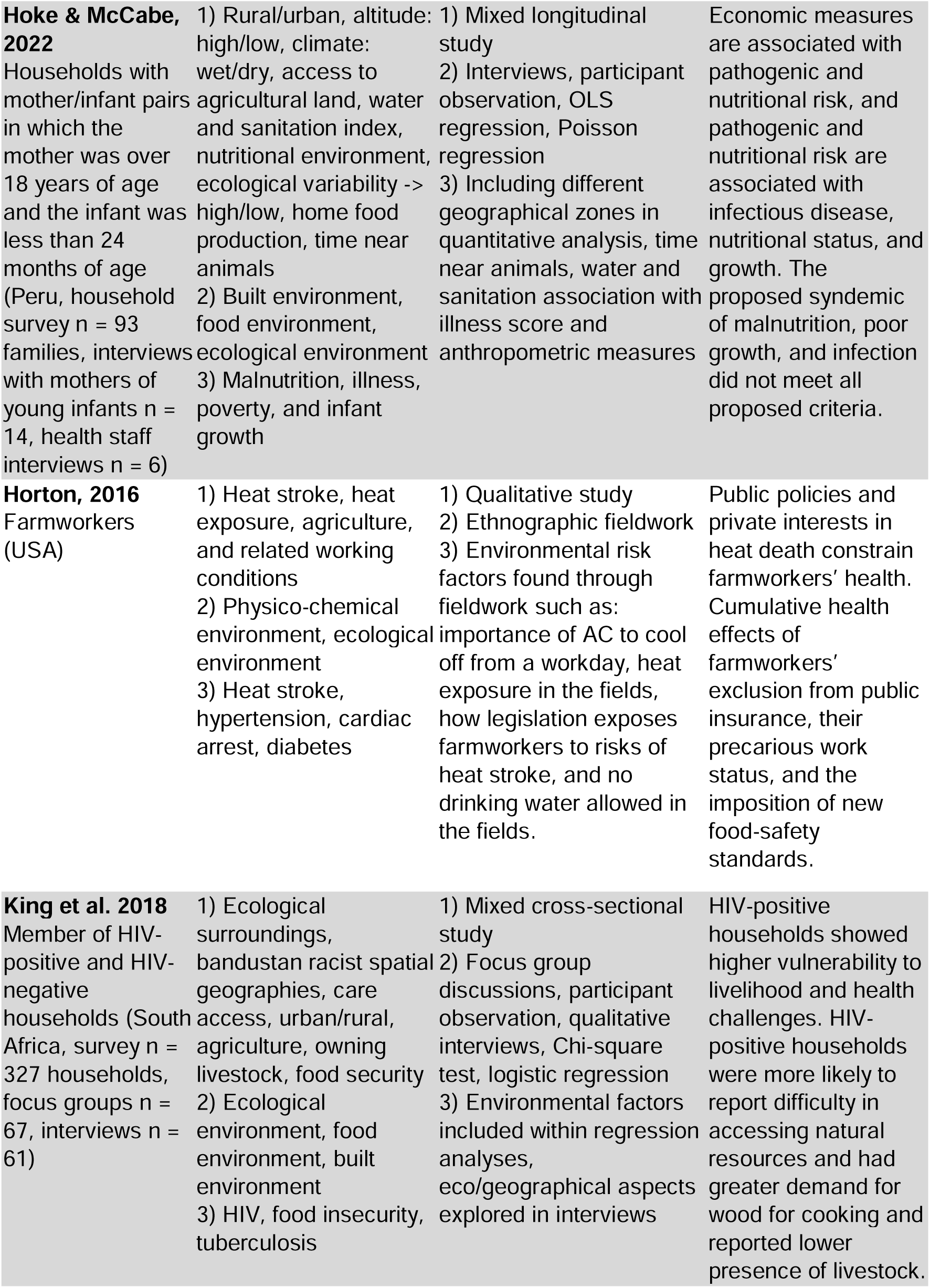
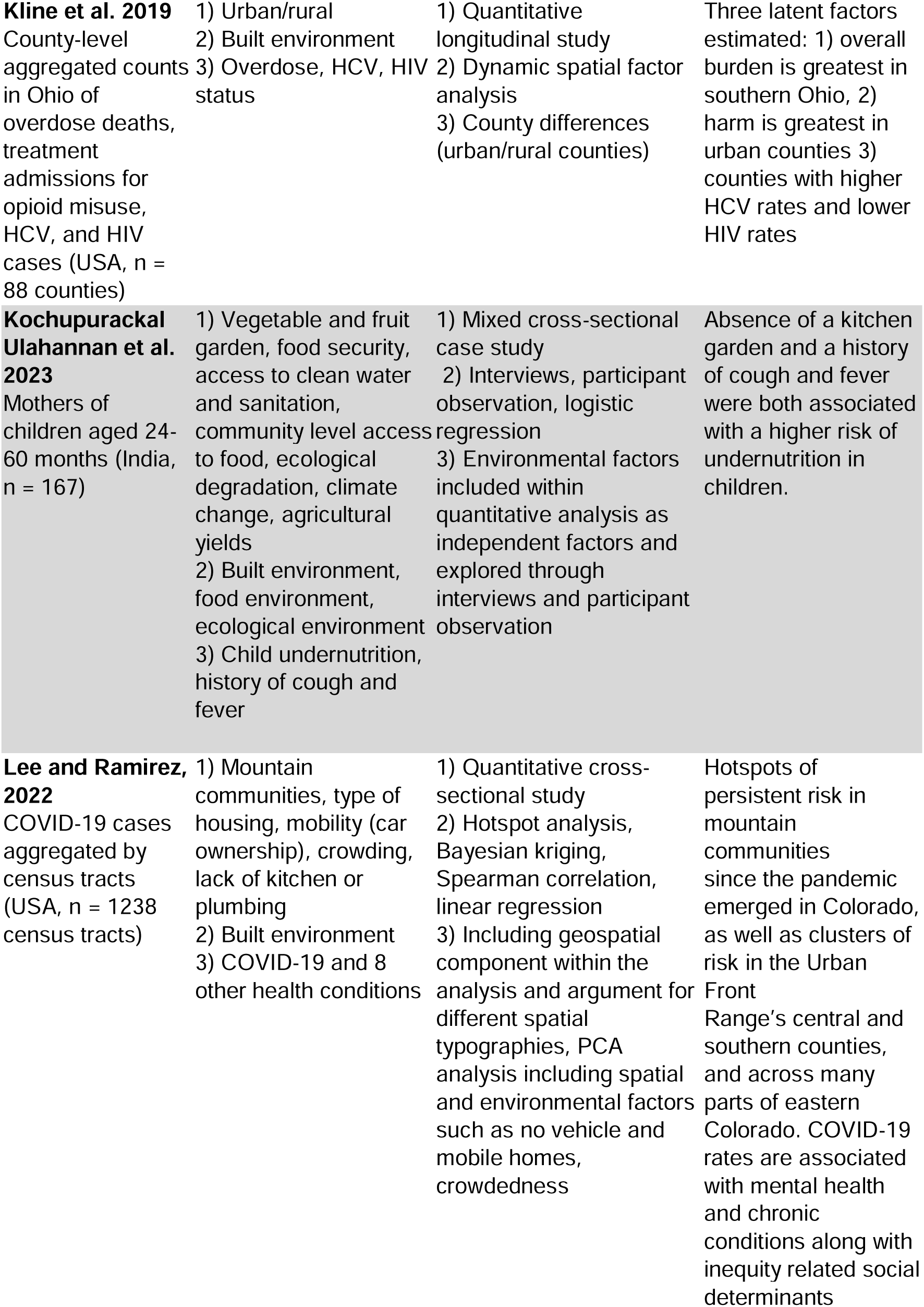
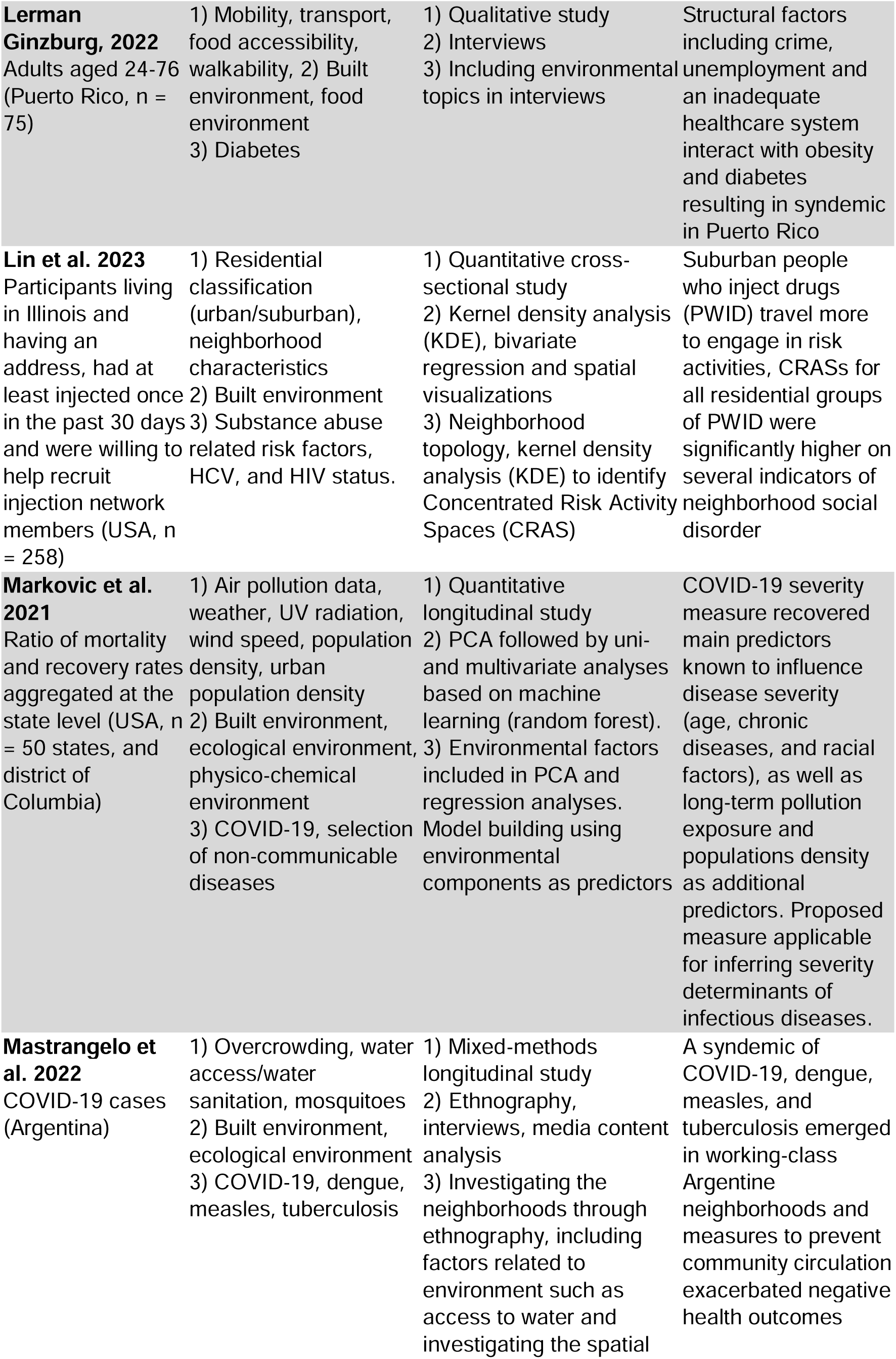
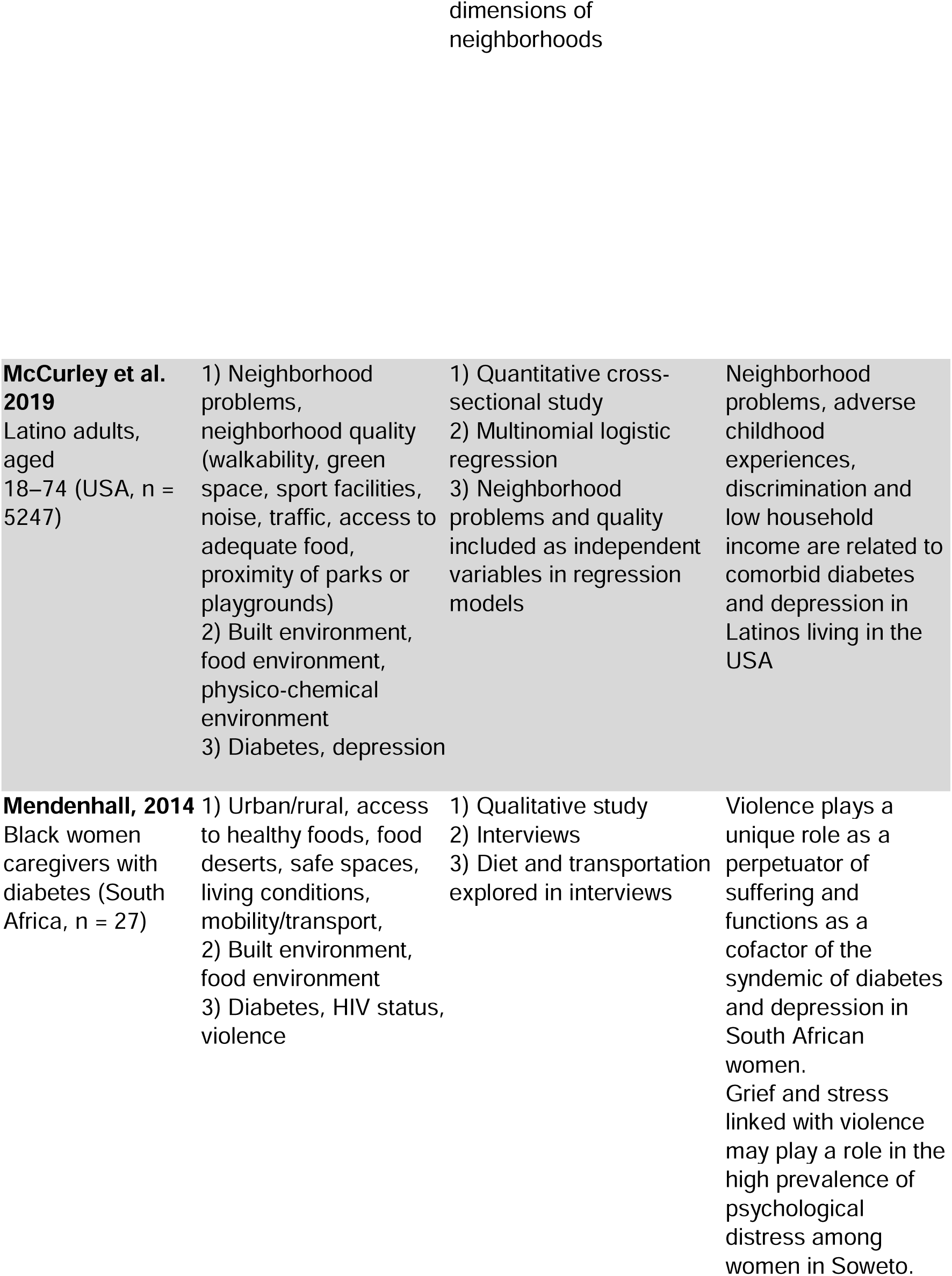
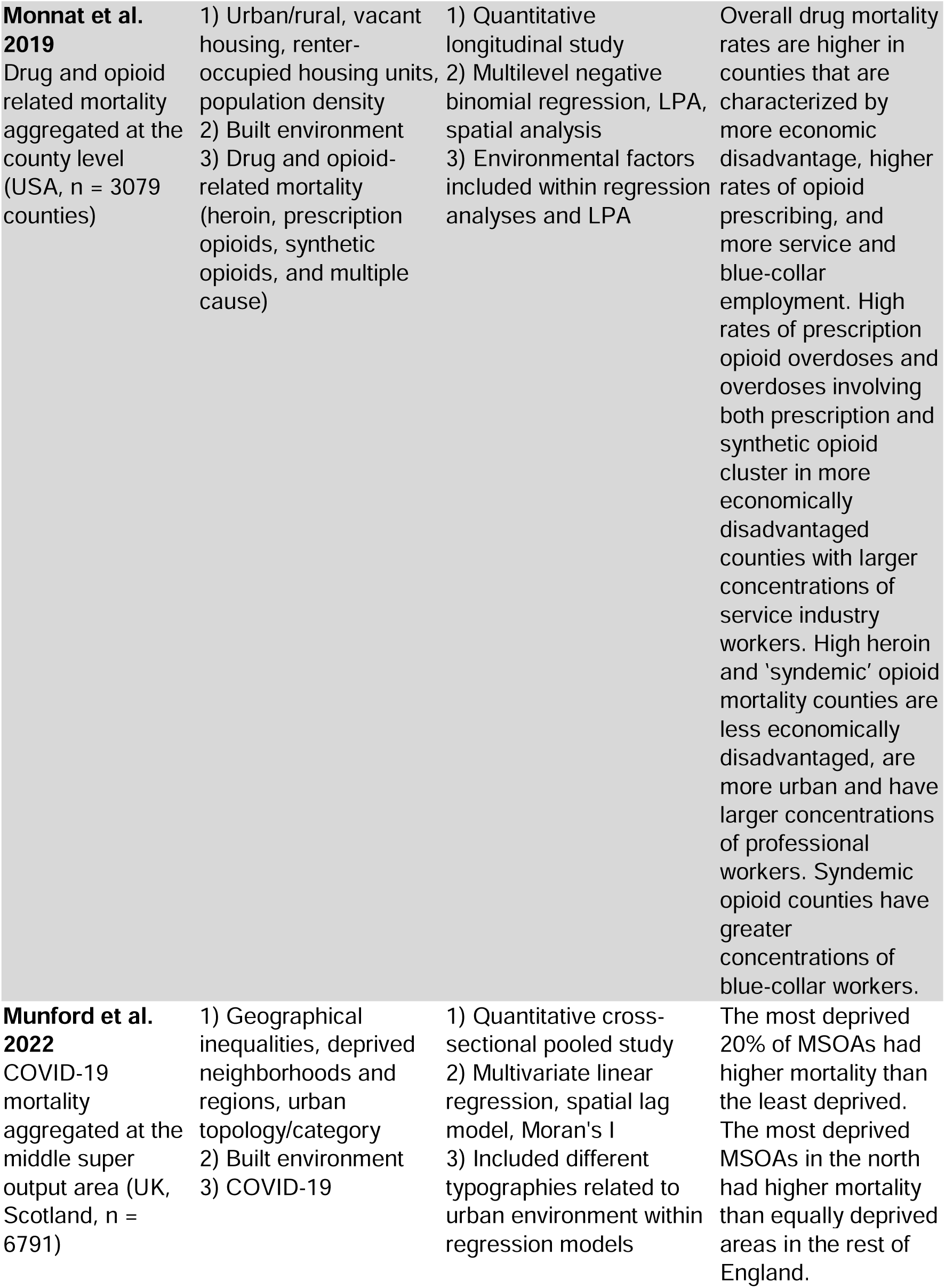
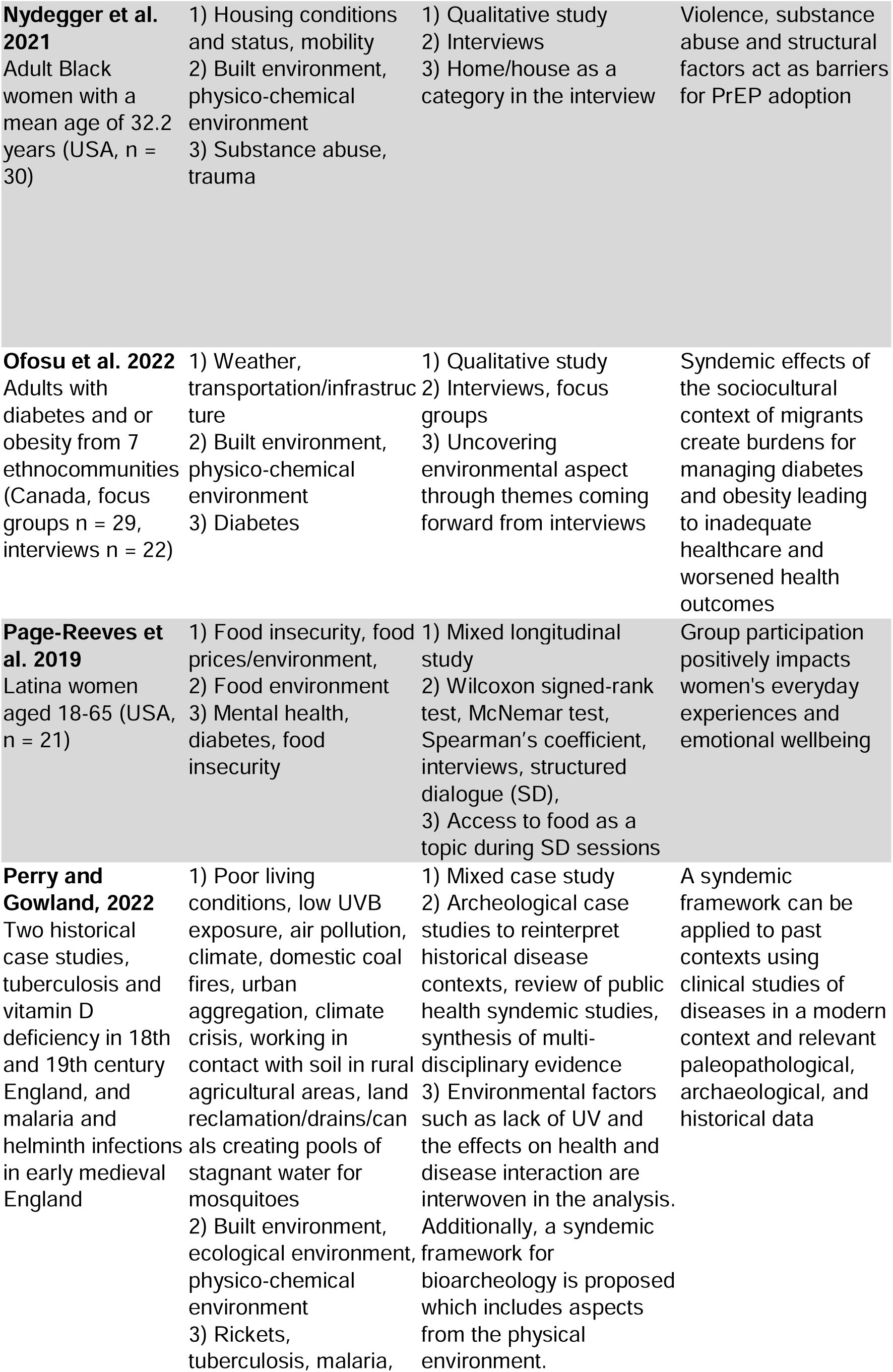
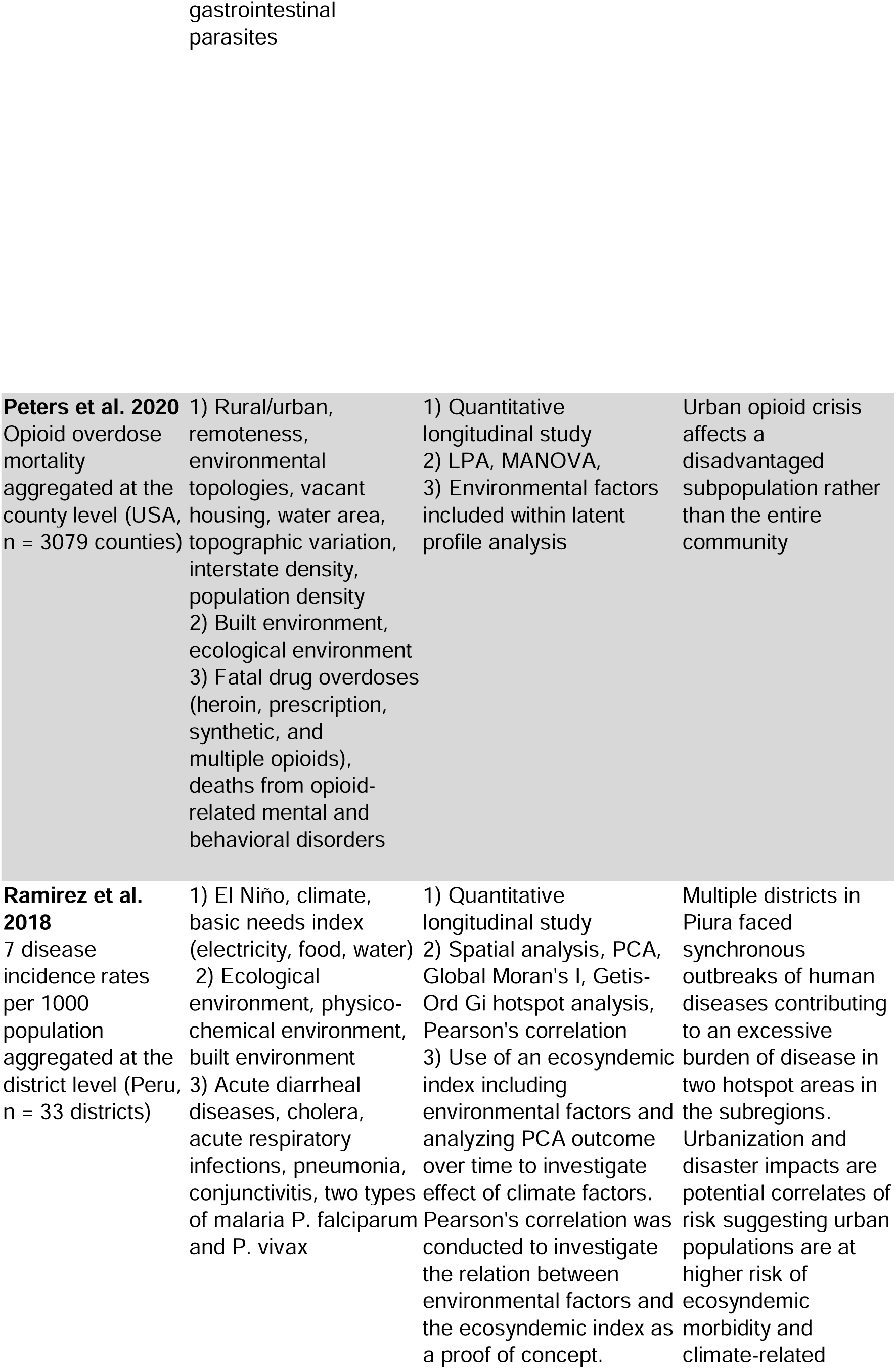
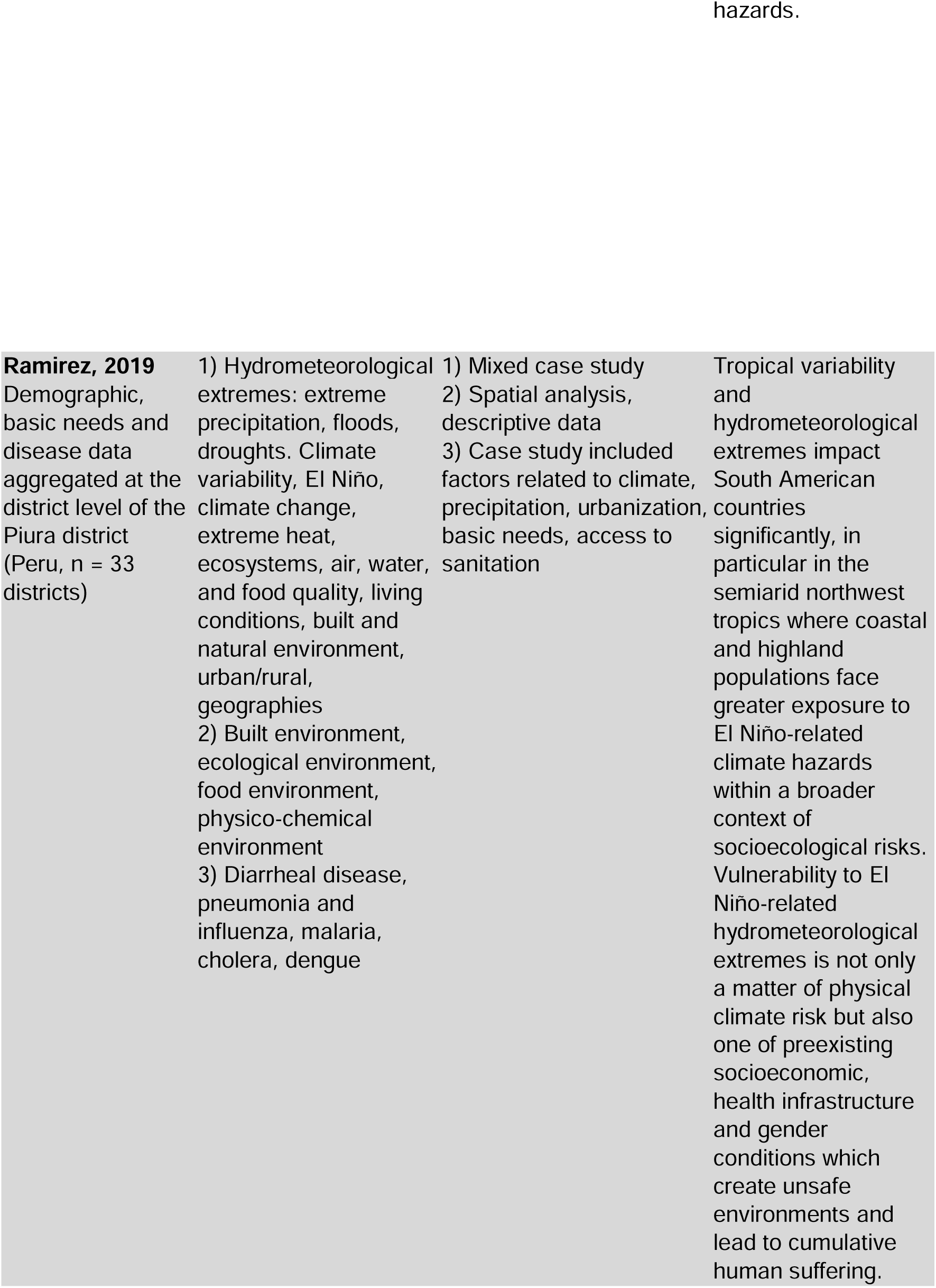
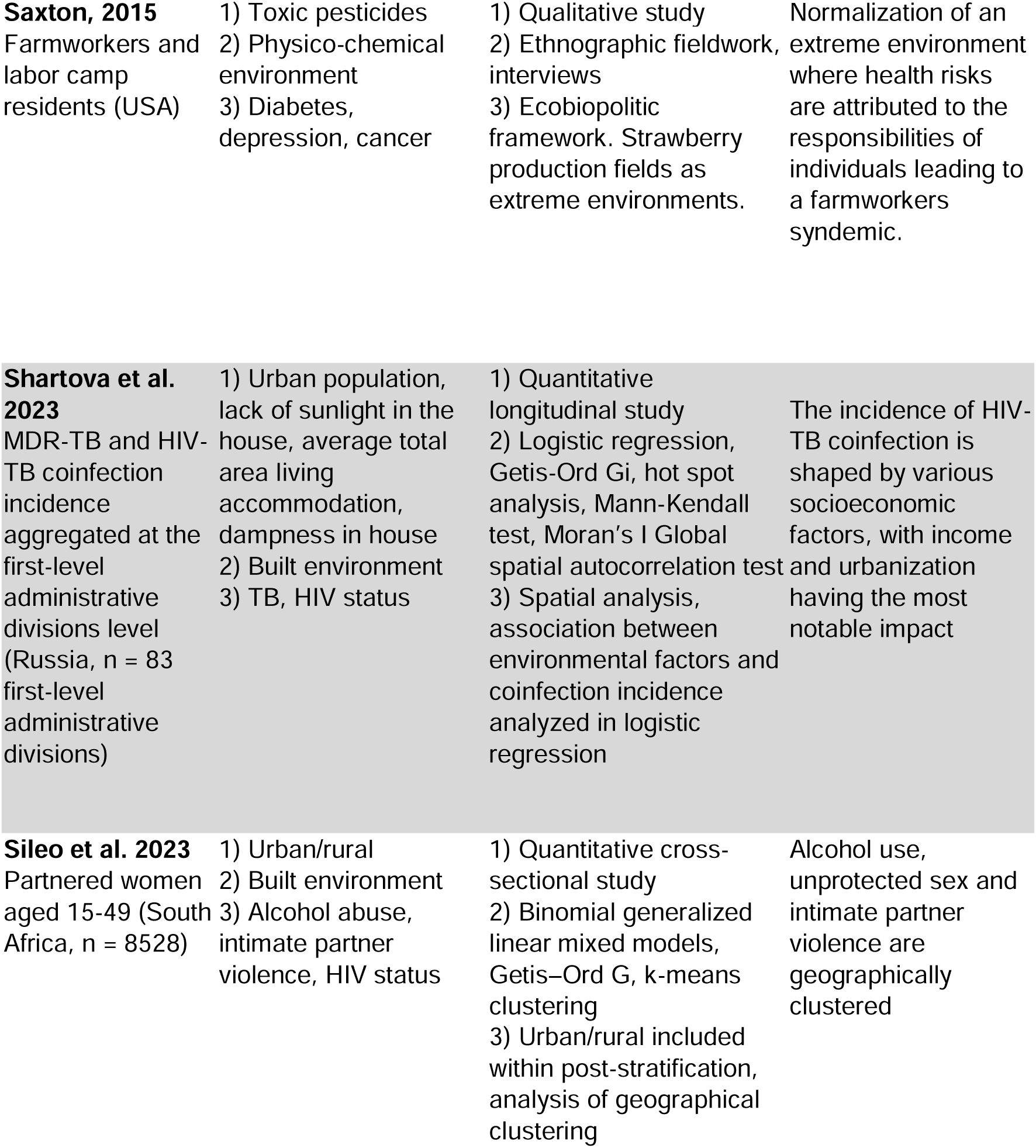
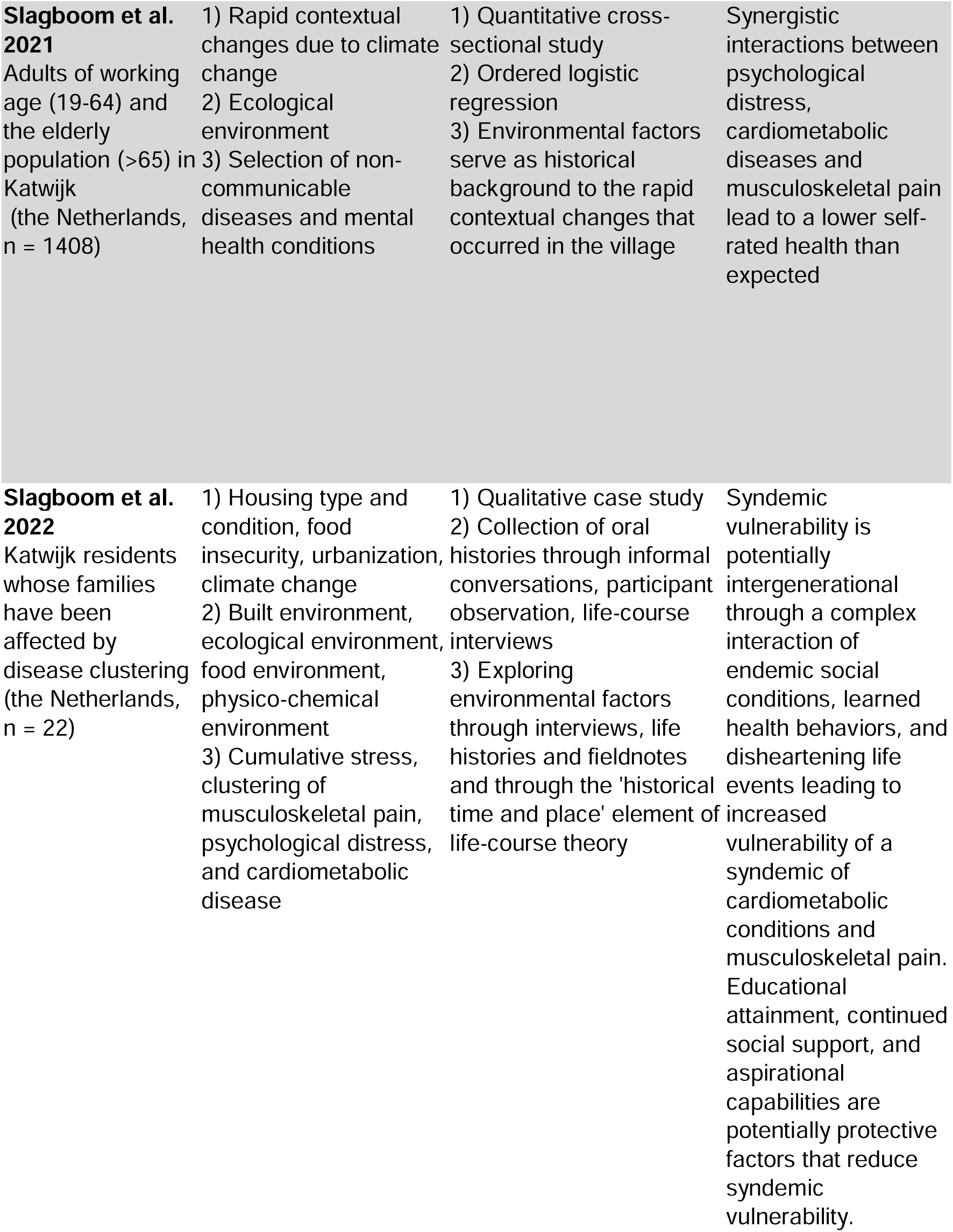
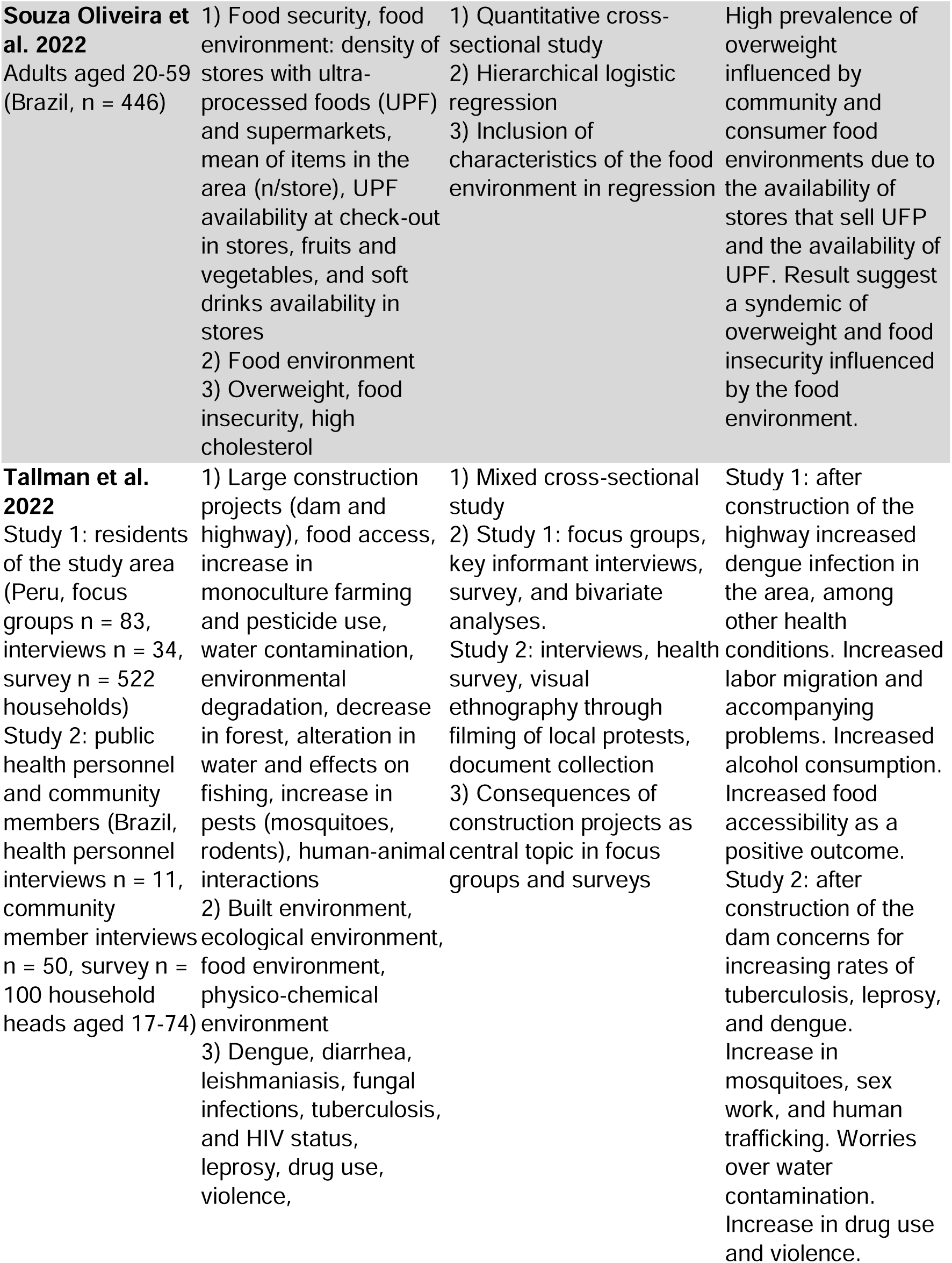
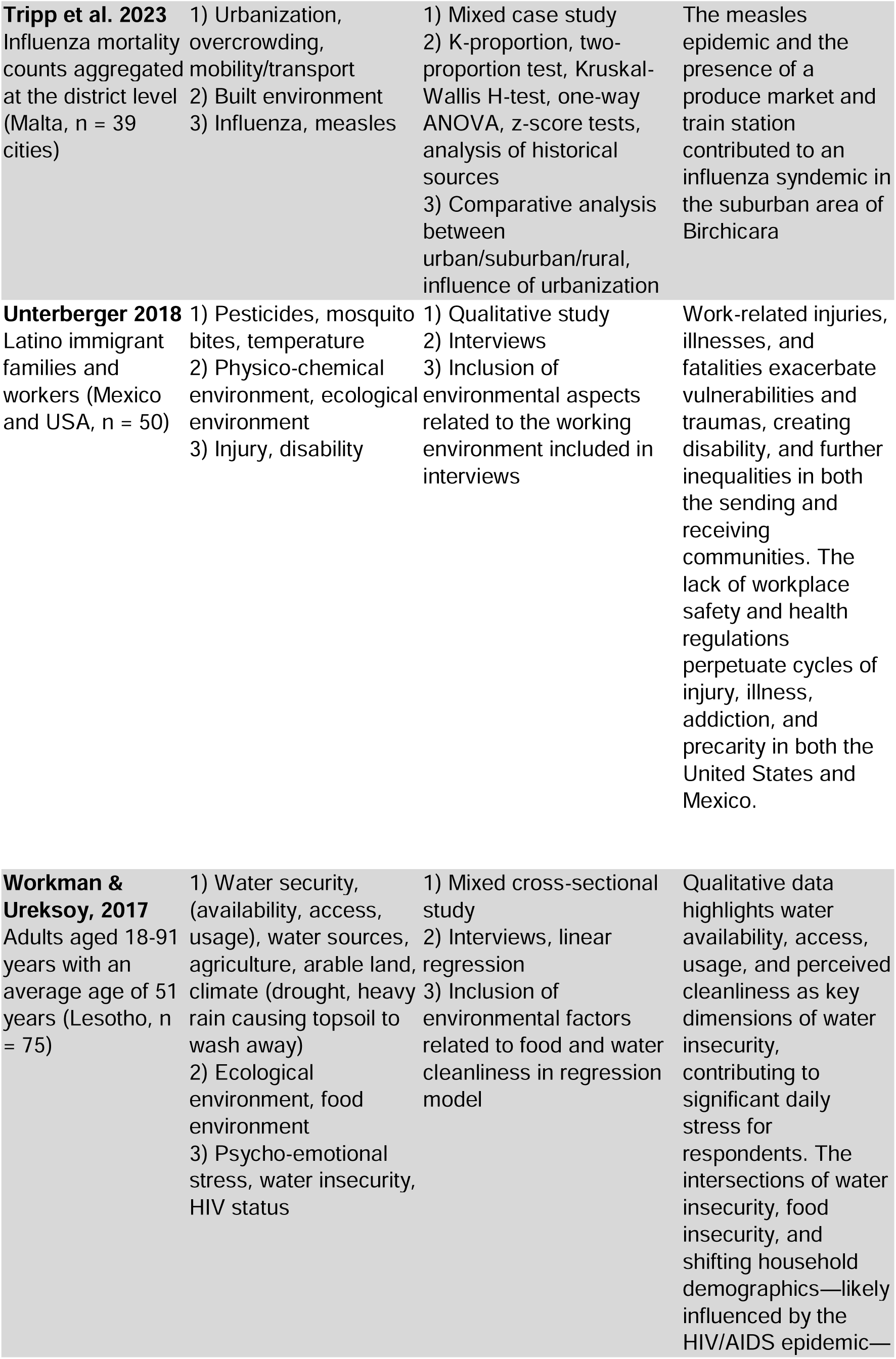
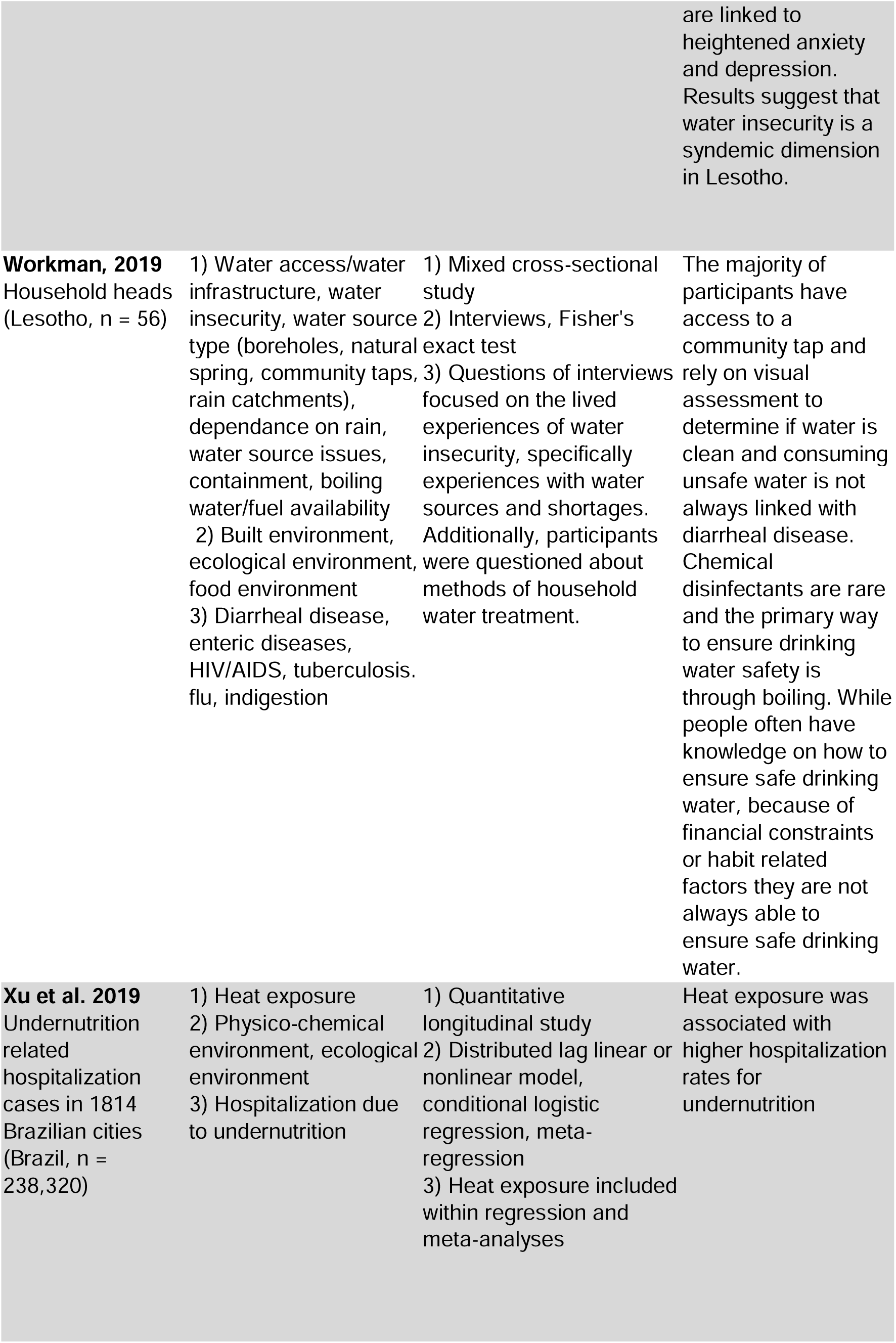
Table of Included Empirical Studies and Their Individual Characteristics (n = 57)

